# Genomic atlas of 7,000 plasma proteins and their associations with diseases and traits in East Asian populations

**DOI:** 10.64898/2026.02.05.26345625

**Authors:** Alfred Pozarickij, Baihan Wang, Ahmed Mohamed, Kuang Lin, Sam Morris, Christiana Kartsonaki, Neil Wright, Hannah Fry, Yiping Chen, Huaidong Du, Derrick Bennett, Ling Yang, Daniel Avery, Dan Valle Schmidt, Liming Li, Jun Lv, Canqing Yu, Dianjianyi Sun, Pei Pei, Junshi Chen, Michael Hill, Richard Peto, Rory Collins, Robert Clarke, Iona Y Millwood, Zhengming Chen, Robin G Walters, China Kadoorie Biobank Collaborative Group

## Abstract

Proteogenomic studies integrating genetic, molecular, and phenotypic data have transformed target discovery, yet remain heavily biased toward European populations. Here, we present a large-scale proteogenomic atlas in a non-European population, analysing 7,289 plasma proteins profiled by SomaScan v4.1 in 3,965 Chinese adults. Genome-wide association analyses identified 3,212 protein quantitative trait loci (pQTLs), including 1,092 proteins with a *cis*-pQTL. Integrating these data with East Asian phenotypes and disease outcomes, we performed proteome-wide phenome scans and identified 7,936 protein-phenotype associations with strong colocalization support (PP.H4 > 0.8). Mendelian randomisation analyses using *cis*-pQTL instruments further prioritised 1,975 protein-phenotype associations, with 645 high-confidence pairs supported by both colocalisation and causal inference. Notably, we identified ancestry-specific pQTLs that contributed to associations undetectable in European studies alone. These associations organised into coherent biological networks, most prominently involving lipid metabolism and cardiovascular disease. Together, this study expands the global proteogenomic landscape and establishes a publicly valuable atlas of genetically anchored protein-phenotype relationships, providing a foundational resource for future genetic, functional, and translational studies, including drug-target prioritisation and risk-benefit assessment.

## Introduction

Proteins play a key role in maintaining the functions of cells, tissues and organs, which include enzymes, antibodies, transport, and structural proteins. Plasma protein levels, whether actively secreted or passively released from healthy and diseased cells, can reflect an individual’s health and disease status. When integrated with genetic and other phenotypic information, plasma proteomics can inform early detection, risk prediction, treatment, and prevention across a wide range of diseases.^1,4^ Recent technological advances in high-throughput proteomic assays now enable cost-effective measurements of plasma levels of thousands of proteins using very small amounts of plasma sample. By integrating genomics and proteomics data in large-scale population-based studies, important insights into the genetic architecture of the plasma proteome can be gained.^5-12^ When combined with genome-wide association studies (GWAS) of clinically relevant phenotypes, protein quantitative trait loci (pQTLs) can be used as instruments in Mendelian Randomisation (MR) and colocalisation analyses to identify putative causal protein-phenotype relationships and to prioritise novel or repurposed drug targets.^13-16^ Recently, two high throughput affinity-based proteomic profiling platforms have been applied to large-scale population-based studies, namely the antibody-based Olink^17^ and the aptamer-based SomaScan platform.^18^ Notably, the Olink Explore platform has been used by the UK Biobank Pharma Proteomics Project (UKB-PPP) to measure 2,923 plasma proteins in 54,219 participants, which led to the discovery of 14,287 pQTL associations.^19^ Similarly, the SomaScan v4 assay including measurements for 4,907 aptamers has been used by deCODE in 35,559 Icelanders, which led to discovery of 18,084 pQTL associations.^9^ The deCODE study also demonstrated that ∼12% of the lead variant-phenotype associations are in high linkage disequilibrium (LD) with at least one SomaScan lead pQTL variant.^16^ Thus, the use of MR to establish causal relationships between proteins and phenotypes provides opportunities to explore the role of proteins in disease risks and aetiology, and identify potential novel therapeutic targets. At least partly due to differences in the assay technology, the pQTLs identified by Olink or SomaScan assays only show moderate replicability in previous studies.^20,24^ Nevertheless both technologies are capable of identifying biologically plausible platform-specific pQTLs, and using both Olink and SomaScan measurements should provide more comprehensive characterisation of the proteomic landscape and its relationship with clinically important phenotypes. Moreover, they also facilitate cross-platform replication and identification of novel associations.

To date, nearly all the reported large proteomic studies have been in European ancestry populations. Our recent analysis of 2,923 Olink proteins in 3,974 Chinese adults identified 1,704 lead pQTL associations, which were fine-mapped to 2,734 credible sets consisting of most likely causal variants.^25^ Approximately 33% of the credible sets did not overlap with the Olink credible sets identified by UKB-PPP, highlighting the added value of pQTL discoveries in diverse ancestries even with much lower sample sizes. However, systematic cross-ancestry evaluation of SomaScan pQTL replicability remains limited. Moreover, the previous deCODE SomaScan GWAS was based on an assay of ∼5,000 aptamers but, with advances in technology, it is now possible to measure up to 11,000 aptamers,^26,27^ providing new opportunities for pQTL discovery. In addition, SomaScan includes aptamers that target multiple proteins, as well as proteins targeted by multiple aptamers, which can offer additional information about protein-protein interactions, different proteoforms and potential epitope effects in which benign missense variants interfere with the assay.

To address the evidence gaps, the present study aimed to 1) characterise the genetic architecture of ∼7,000 proteins measured with the SomaScan v4.1 assay in 3,965 Chinese participants from the China Kadoorie Biobank; 2) create a comprehensive atlas of potential causal protein–trait relationships between using Mendelian randomisation (MR) and colocalisation analyses; and 3) cluster protein-trait relationships into biologically coherent networks to prioritise proteins for therapeutic targeting and prevention strategies.

## Results

### Baseline characteristics and study design

The baseline characteristics of the 3,976 study participants are summarised in **Supplementary Table 1**. The mean age of participants was 60.3 years (SD 11.5), and 48.8% were from urban regions. Participants had been selected for an ischemic heart disease (IHD) case-subcohort study, and those with IHD were on average ∼12 years older than those in the sub-cohort, had a lower proportion of females (45% vs 62%), had a higher prevalence of hypertension and diabetes, and had higher mean systolic blood pressure (SBP) and body mass index (BMI), and lower physical activity levels (median 13.0 vs 21.3 MET hours/week). **Supplementary Figure 1** provides an overview of the study design, analytical strategy, and main findings.

### Association of genetic variants with plasma protein levels

*We* performed genome-wide association studies (GWAS) of ∼11 million imputed variants (minor allele count > 20, MAP > 0.0025) for 7,596 aptamers **(Supplementary Table 2)**, using the ANML-normalised version of SomaScan v4.1 data (see Methods). The GWAS showed minimal inflation (mean genomic λ_gc_ - 1.003, range 0.979-1.027), indicating appropriate control of population structure and technical artefacts. We have previously shown that including IHD cases in our study design does not bias the effect sizes of the associated genetic variants.^25^ At a Bonferroni corrected significance threshold *(p* < 5 x 10 ^-9^/7,596), 3,212 pQTL associations comprising 1,282 *cis*-pQTLs, 1,825 *trans*-pQTLs and 105 associations with nonhuman targets were identified **(Figure 1A; Supplementary Table 3)**. Overall, 2,427 (32%) human aptamers, corresponding to 2,187 unique proteins had pQTLs, of which 938 aptamers had only *cis*, 1,159 had only *trans*, and 330 had both *cis* and *trans* pQTLs. The median number of pQTL signals per aptamer was 1 (range 1-4). Notably, 179 aptamers had strong *trans*-pQTLs with *p* < 1 x 10^100^ yet had no *cis*-pQTL for the target protein. The 3,212 pQTLs were mapped to 715 non-overlapping genomic regions, of which 475 regions included only *cis*, 79 only *trans*, and 161 both *cis* and *trans* associations **(Supplementary Table 4)**. The number of protein associations per locus ranged from 1 to 185 (median = 1). There were 328 loci associated with more than one protein, and 41 highly pleiotropic loci associated with >10 distinct proteins. Many of the most pleiotropic loci corresponded to well-known regulators of plasma proteins, such as the ABO blood group locus, APOE/APOC1 locus, and the human leukocyte antigen (HLA) region. Among the 105 (of which 102 mouse-related) non-human associations targeted by 86 aptamers, 97% were mapped to a single immunoglobulin heavy chain locus (IgH; UniProtID=Q99LC4) in the genome (spanning immunoglobulin gene clusters), with ∼30% sharing the same lead SNP. This suggests that many of the aptamers designed for non-human targets (for instance, mouse IgH orthologs or other control antigens) are picking up a common genetic effect, likely related to immunoglobulin levels or assay background.

**Figure 1.**
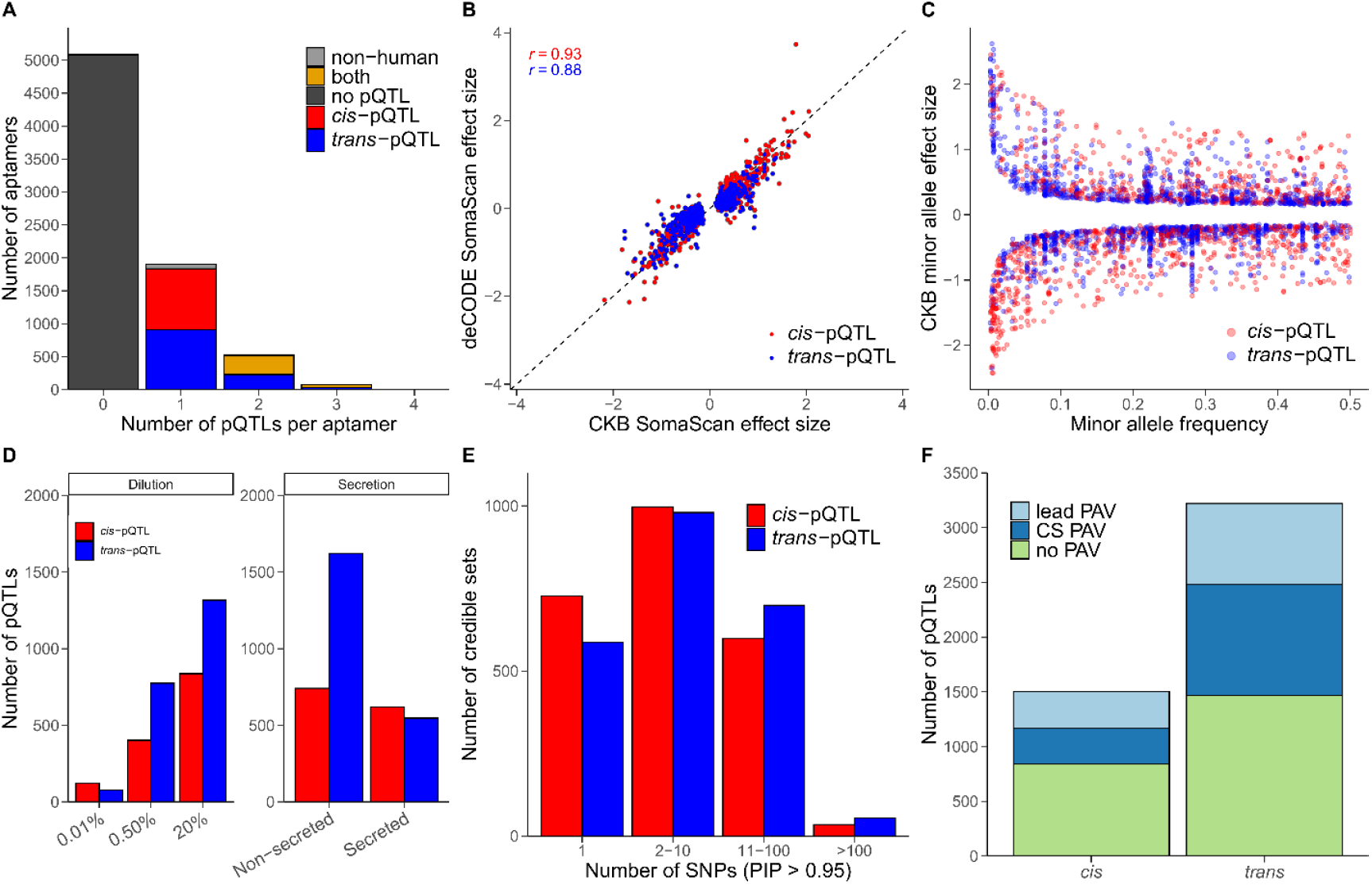
Overview of pQTL associations with plasma protein levels. (A) Distribution of the pQTL associations (*P* < 6.58 x 10^-13^) per SomaScan aptamer (red: *cis;* blue: *trans)*. (B) Comparison of lead CKB pQTL effect sizes in deCODE. (C) Minor-allele effect sizes of CKB pQTLs as a function of minor allele frequency. (D) Distribution of pQTL counts stratified by SomaScan protein annotations. (E) Distribution of the number of causal SNPs within fine-mapped credible sets. (F) Annotation of *cis-* and *trans*-pQTLs according to whether the sentinel pQTL was protein-altering (lead PAV) or whether a protein-altering variant for the target protein was present within the fine-mapped credible set (CS PAV).

### Comparison of pQTLs across ancestries and platforms

We compared pQTL associations in three independent datasets: (i) the deCODE study of ∼35,000 Icelanders (the SomaScan v4.0 assay of 4,907 aptamers),^9^ (ii) the UKB-PPP of ∼55,000 participants (the Olink Explore platform of 2,973 proteins),^19^ and (iii) a CKB study of 3,976 participants (the Olink Explore platform of 2,973 proteins) as the present study.^25^ For the Olink datasets, comparisons were made for 2,168 overlapping proteins captured by both platforms. Despite differences in ancestry and technology, a high proportion of pQTL signals showed consistent effect magnitude and direction. Overall, 87% of lead pQTLs identified in this study were replicated in the deCODE SomaScan at *p <* 5 x 10 ^-8^ with concordant effect directions (94% had nominal replication at p < 0.05; **Figure 1B; Supplementary Table 5)**. Replication rates in deCODE were slightly higher for *cis*-pQTLs than for *trans*-pQTLs (92% vs 84% at *p* < 5 x I0^-8^; 96% vs 92% at *p* < 0.05), consistent with *cis*-acting variants generally having larger effect sizes.^19^ The effect size estimates between CKB (East Asians) and deCODE (Europeans) were highly correlated (Pearson r = 0.93 for cis-pQTL effect sizes and r = 0.88 for *trans*-pQTLs), suggesting that the vast majority of lead pQTL effects on protein levels are consistent across different ancestry populations. In the CKB Olink dataset, >80% of lead pQTLs were replicated with higher replication rates observed for *cis*-pQTLs than for *trans*-pQTLs (92% vs. 83% at *p* < 5 x 10 ^-8^). There were only moderate effect size correlations between SomaScan and Olink pQTLs **(Supplementary Figure 3)**. Only 65% of the CKB lead pQTLs could be replicated in the UKB-PPP Olink dataset at *p <* 5 x 10 ^-8^ with concordant effect direction, again with higher replication rate for *cis*-pQTLs than for *trans*-pQTLs (83% vs 44% at *p <* 5 x 1087% ; ^-8^ vs 53% at p < 0.05). The discrepancy between *cis* and *trans* pQTL replication may reflect the smaller effect sizes and context-dependence of many *trans*-pQTLs, as well as potential differences in the epitope or protein isoform captured by the two assays. In total, 25% of lead pQTLs were replicated in all three datasets with *p <* 0.05 and concordant effect direction.

### Characteristics of pQTLs

As has been previously observed^7,19,25^, pQTLs with lower allele frequency tended to have larger effects on protein levels **(Figure 1C)**. The effect size estimates (change in protein level in standard deviations per effect allele) ranged from -2.43 to +2.62. Overall, *cis*-pQTLs had significantly larger effects than *trans*-pQTLs (median absolute β = 0.21 for *cis* vs 0.16 for *trans;* Wilcoxon P⍰=l⍰13.5 x 10 ^-20^). We identified 149 pQTLs with MAF < 1%, of which 67 were *cis*-pQTLs. The proportion of protein level variance explained by each sentinel pQTL ranged from 0.01% up to 76% **(Supplementary Table 3)**. As expected, *cis*-pQTLs tended to explain more variance in their target protein levels than *trans*-pQTLs (median r^2^ - 3.8% for *cis* vs 1.6% for trans). Some *cis*-pQTLs had exceptionally large r^2^ values (>70%), particularly those associated with *ABO, LILRA3*, and *SIGLEC9*, which often reflect protein-altering variants or gene deletions that dramatically impact the protein levels or function. All *cis*-pQTLs were strong instrumental variables (F-statistic > 27). We assessed whether pQTL discovery depends on protein abundance and secretion. There was a clear trend for more abundant proteins (those measured at higher dilution in the assay) to have more genetic associations. Roughly, each additional dilution step (approximately corresponding to a 2-fold decrease in protein abundance in the sample) was associated with double the number of pQTLs, for both *cis* (0.01%=120; 0.5%=403; 20%=839) and *trans* (0.01%=77; 0.5%=774; 20%=1,316) signals **(Figure 1D)**. Of the 2,322 proteins with at least one pQTL, 29% were secreted into blood. Secreted proteins where equally likely to have *cis* and *trans* pQTLs (619 vs 547) unlike nonsecreted proteins with much greater number of *trans* compared to *cis* (741 vs 1620). One speculative explanation is that many non-secreted proteins in plasma are indirect biomarkers of broader processes (hence more *trans* influences), whereas secreted proteins (often hormones or cytokines) more directly influenced by genetic variation at the locus encoding that protein.

### Fine-mapping and functional annotation of pQTLs

To identify causal variants, we performed Bayesian fine-mapping for each pQTL locus. At 95% posterior probability, we obtained credible sets (CS) for 3,160 pQTLs (>98% of all pQTLs), containing a total of 66,333 candidate causal SNPs across all sets **(Supplementary Table 6-7)**. In total, 4,905 distinct CS were identified. The median number of CS per pQTL was 1 for both *cis* (max=10) and *trans* (max=9). Notably, 666 *cis*-pQTL signals and 1,471 *trans*-pQTL signals were fine-mapped to single-SNP resolution **(Figure 1E)** and *cis*-pQTLs were generally fine-mapped more confidently: the lead variant’s posterior inclusion probability (PIP) was higher on average for *cis* loci (median PIP ∼53%) than *trans* loci (median PIP ∼45%). In ∼70% of cases where a CS was identified, the lead pQTL was also the highest-PIP variant in the set for both *cis*-pQTLs (73%) and *trans*-pQTLs (69%). In a further ∼20% of cases, the sentinel SNP was included in the CS but was not the top causal candidate SNP. This suggests that potentially 10% of loci represent complex combinations of overlapping signals in which a non-causal variant becomes the lead signals due to LD. We next functionally annotated the fine-mapped variants **(Supplementary Table 6)**. Among *cis-* pQTLs, ∼47% of the likely causal variants were intronic and 15% were intergenic regulatory variants, while 18% were missense mutations (non-synonymous coding variants). Only ∼1% of *cis*-pQTLs were caused by deleterious coding variants (e.g., 13 stop-gained mutations, 6 in-frame insertions/deletions, 3 start-loss). For *trans-*pQTLs, the most common annotations were intronic (41%), missense (27%), and intergenic (15%). However, *trans*-pQTLs had fewer pQTLs with deleterious consequences. We defined protein-altering variants (PAVs) as coding variants predicted to alter the protein sequence (e.g., missense, splice acceptors/donors, start lost, stop gained/lost, frameshift, in-frame deletion/insertion). Among *cis*-pQTLs, 483 fine-mapped top SNPs were PAV, and an additional 243 pQTLs had at least one PAV for the target protein in their 95% CS **(Figure 1F)**. Thus, roughly 30% of *cis*-pQTLs potentially involve a coding change in the protein itself. For *trans*-pQTLs, 704 (28%) CS SNPs contained PAV. These likely represent cases where a coding variant in protein X leads to altered expression of gene Y (for example, by altering binding to an enhancer or other transcriptional regulation site). Our findings confirmed that a substantial fraction of pQTLs can be directly tied to protein-coding changes in the genome, whereas the remainder are presumed non-coding regulatory variants.

### Protein-protein network

*We* identified 305 proteins for which a genetic variant modulated the circulating level of its cis-target protein and, in turn, exerted downstream effects on 1,018 *trans*-protein targets **(Figure 2A)**. Approximately 64% of *trans*-proteins exhibited effects in the same direction as the *cis*-protein, 35% in the opposite direction, and 1% in both directions. The latter pattern arose in situations where multiple SomaScan aptamers targeted the same protein, such that one aptamer captured a positive association and another a negative association. This was observed for seven proteins (APOE, CCL23, CFHR5, FCN3, SERPINA10, SERPING1 and TNC). Notably, the effect-size distributions for *trans*-proteins with concordant and discordant directions were highly similar, with comparable absolute median β estimates (0.25 versus 0.24, respectively). Of the 305 proteins, 167 exerted downstream effects on more than one *trans* protein. The most pleiotropic signals were observed at the SDF2 and VTN locus on chromosome 17 (chr17:27-30 Mb), each influencing 161 *trans*-proteins, followed by the apolipoproteins APOE, APOC3 and APOA5, which affected 122, 78 and 76 *trans*-proteins, respectively **(Figure 2B, Supplementary Table 8)**. The *cis-trans* regulatory network showed a highly ordered architecture. Whereas numerous proteins formed sparse connections, a comparatively smaller subset of hub proteins organised densely interconnected clusters, thereby underscoring a restricted group of secreted proteins with broad influence across the plasma proteome. In addition to apolipoproteins, prominent hubs included complement component and regulator proteins (CFH, CFHR1, C1R, C1S, C4A/C4B, CFB), coagulation and kinin pathway proteins (F5, KNG1, HRG, VTN, SERPINA10), and carrier or scaffold proteins (BCHE, ABO, ITIH1, OBP2B, C0LEC11). Among the proteins with *cis*-pQTLs that also had downstream *trans*-effects, the median absolute observational correlation was more than twofold higher than among proteins without a *cis-trans* relationship (median |r| = 0.157 vs 0.075). For proteins in which *cis* and *trans* pQTL effects were concordant in direction, the median correlation was 0.21, compared with -0.01 for proteins with discordant pQTL effects. We also identified 113 *trans*-proteins which were influenced by multiple *cis*-proteins. Notably, METTL2B, a methyltransferase-like protein that is involved in the methylation of tRNA was influenced by 10 proteins all located in the HLA region.

**Figure 2.**
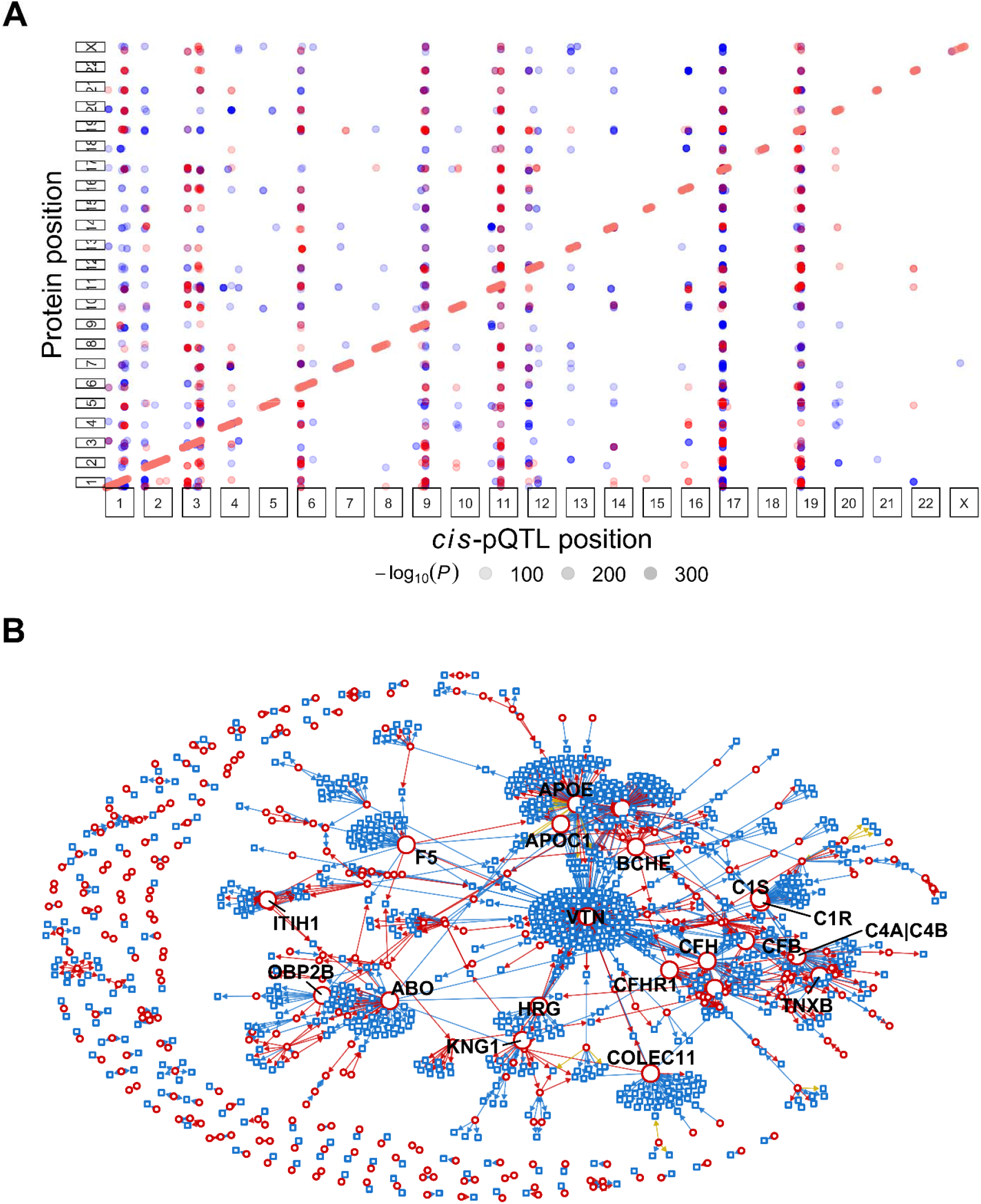
Integrative view of the *cis*-pQTL genomic architecture and the resulting *trans*-protein regulatory network. (A) Genomic positions of *cis*-pQTLs (diagonal clusters; *P <* 6.58 x 10 ^-13^) that also exert *trans*-effects on other proteins. Red points indicate positive *trans*-effects, whereas blue points indicate negative *trans*-effects, and point transparency reflects the strength of association. (B) Network representation of *trans* -protein regulatory interactions driven by *cis*-acting pQTLs. Red circles indicate proteins with *cis*-pQTLs, blue squares denote proteins that are influenced through *trans* -effects exerted by *cis*-regulated proteins. *Trans*-effects are coloured by direction: red for positive, blue for negative, and gold for bidirectional effects resulting from multiple aptamers recognising the same protein.

### Overview of traits across three East-Asian biobanks

*We* obtained East-Asian GWAS summary statistics for 274 traits to perform phenome-wide scans of the pQTLs. After accounting for overlapping traits between biobanks, there were 225 unique phenotypes **(Supplementary Table 9)**, comprising 100 quantitative traits (43 biomarkers, 37 lifestyle factors, 17 physical measurements, and 3 reproductive traits) and 125 binary traits (98 disease outcomes, 22 medication-use phenotypes, 2 lifestyle, 1 reproductive, and 2 miscellaneous traits). For disease and medication use traits, we required each phenotype to have at least 500 cases in GW AS. The binary trait with the smallest case number was polycystic kidney disease (510 cases), while the largest was smoking status (ever vs never smoker, 88,277 cases). The majority of the GWAS data came from BioBank Japan (BBJ; 179 traits, 65%), with 60 traits (22%) from the Korean Genome and Epidemiology Study (K0GES) and 35 traits (13%) from Taiwan Biobank (TWB did not have publicly available GWAS for binary traits, so its contribution was limited to quantitative traits). There were 18 phenotypes measured in all three biobanks and 11 phenotypes present in two of the biobanks. Notably, 19 phenotypes including cardiometabolic traits such as blood pressure and blood lipid levels were represented by all biobanks, allowing cross-biobank comparisons.

### Phenome-wide associations of pQTLs

*We* confirmed that the effect allele frequencies of the lead pQTL variants identified in this study were highly consistent with those reported in external EAS biobanks (Pearson r≈0.98; **Supplementary Figure 4)**. A small number of variants showed noticeable frequency discrepancies, potentially reflecting technical issues such as strand alignment errors. However, there was no evidence of systematic bias, suggesting that these differences more likely reflect population-specific allele frequency variation. We performed a phenome-wide association study (PheWAS) across 3,107 lead pQTLs (p < 6.58 x 10 ^-13^) targeting human proteins and 225 traits. In total, we identified 17,980 protein-trait associations at FDR < 0.05 **(Supplementary Table 10)**, comprising 1,650 proteins and 223 traits. Of these associations, 16% were *cis*-acting and 84% were *trans*-acting. Among proteins that passed FDR threshold, 513 had *cis*-only associations, 973 had *trans*-only and 164 had both *cis* and *trans*. Approximately 19% of all protein-trait associations involved disease outcomes, and one-third of these were concentrated in five major cardiometabolic conditions: hyperlipidaemia (n = 306), myocardial infarction (268), coronary artery disease (244), stable angina pectoris (216) and type 2 diabetes (139). Hierarchical trait clustering highlighted several coherent groups of traits with similar protein-association profiles **(Supplementary Figures 5-8)**. One of the most prominent clusters grouped LDL-C and total cholesterol biomarkers with coronary artery disease, stable angina pectoris and related medication-use traits, including HMG-C0A reductase inhibitors, vasodilators used in cardiac disease and salicylic acid derivatives **(Supplementary Figure 8C)**. In total, 746 proteins with FDR < 0.05 were associated with at least one trait in this cluster, 211 of which were associated with five or more traits; notably, six proteins (APOB, GRIPI, EMC4, ITM2B, PCSK9 and TEK) were associated with all traits in the cluster. A closely related cluster comprised blood pressure (BP) lowering medications (β-blocking agents, calcium-channel blockers and agents acting on the renin-angiotensin system), whereas all quantitative blood pressure traits (systolic, diastolic, mean arterial and pulse pressure) formed a distinct cluster characterised by associations with 365 proteins (305 of which overlapped with the BP medication cluster). Among these, AGT, ALDH2, ARL3, ENPEP and SCUBE1 were associated with all four blood pressure traits. Hypertension showed a protein-association profile more similar to intracerebral haemorrhage; however, this separation likely reflects limited power in the underlying GWAS rather than a biologically distinct pathway. Another large cluster comprised vitamin intake measures and the use of nutritional supplements such as zinc, folate and calcium. Type 2 diabetes clustered closely with diabetes medications, glucose intake, fasting glucose and HbA1c, forming a metabolic cluster that included 327 associated proteins, of which CPB1, MLN, PCLAF, PNLIPRP1 and PRSS2 were associated with all traits. With the exception of waist-to-hip ratio, which clustered with glaucoma and antiglaucoma preparations, and height, which showed the closest similarity to forced vital capacity and forced expiratory flow, anthropometric traits (body fat rate, body mass index, weight and hip/waist circumference) clustered together. Multiple tightly linked immune-related clusters were also observed. For example, rheumatoid arthritis, immunosuppressant use and non-steroidal antiinflammatory and antirheumatic drug use clustered with osteoporosis and medications affecting bone structure and mineralisation; these traits were further closely linked to endocrine disorders, including hypothyroidism, hyperthyroidism, thyroid hormone preparations, Graves’ disease and Hashimoto’s disease. Respiratory traits formed a separate cluster in which adrenergic inhalants, asthma, chronic obstructive pulmonary disease and the use of glucocorticoid medication clustered together, whereas allergic rhinitis, pollinosis, allergic conjunctivitis, the use of antihistamine medication and atopic dermatitis formed a different cluster that was more closely related to chronic sinusitis and varicella-zoster or herpes infections cluster.

### Colocalisation and Mendelian randomisation analyses

*We* identified 7,936 protein-trait pairs with strong evidence of colocalisation (PP.H4 > 0.8) across 1,036 proteins and 162 traits, including 2,169 pairs with very high support (PP.H4 > 0.99; **Supplementary Table 10; Supplementary Figure 9)**. These findings indicate substantial sparsity of shared causal variants, as fewer than 2% of all possible protein-trait pairs showed evidence of colocalisation. Notably, most shared signals (91%) were located in *trans* regions. Among *cis*-pQTLs, shared signals were more frequent when the lead variant was located within the structural gene boundaries than when it was located in flanking regulatory region (451 versus 249 colocalisations). Restricting clustering to the 98 disease outcomes **(Figure 3)**, which accounted for nearly 20% of all colocalised signals (1,536 pairs), revealed 423 proteins sharing a signal with 62 diseases. Circulatory diseases, such as coronary artery disease, aortic aneurism, peripheral arterial disease, showed the greatest number of colocalisations (259 proteins and 14 diseases), followed by metabolic diseases such as type 2 diabetes and thyroid-related conditions (293 proteins and 7 diseases; **Supplementary Figure 11)**. The top three proteins, ALDH2, AGT, and SCUBE1, colocalised with 22-23 conditions, with only ALDH2 having a *cis*-pQTL. For non-disease traits, we identified 6,400 protein-trait colocalising pairs involving 1,003 proteins and 100 traits **(Figure 4)**. These pairs were dominated by molecular and physiological phenotypes: biomarker measurements accounted for 1,715 colocalisations, followed by blood-cell traits (1,463), protein biomarkers (988), drug-exposure phenotypes (728), enzyme activities (679), and anthropometric traits (458). Circulatory, dietary, reproductive, respiratory, alcohol-related, metabolic, and smoking-associated traits were also represented, reflecting a diverse yet uneven distribution of shared genetic signals across biological domains. A limited subset of proteins displayed extensive pleiotropy, most notably ALDH2, SCUBE1, and AGT, which colocalised with 54, 52, and 51 traits, respectively **(Supplementary Figure 12)**. These far exceeded the next tier of proteins, including PCLAF and GOLM2 (30 colocalisations each), GOLM1 (29), QSOX2 and STAR (27 each), and PLXDC2 and MEP1A (26 each). Combining results across all traits **(Supplementary Figure 9)** provided a more comprehensive view of the global protein-trait colocalisation landscape and revealed broad, partially overlapping trait groupings that qualitatively resembled clusters previously identified by PheWAS, including cardiovascular, anthropometric, and blood pressure-related traits. However, quantitative comparison of cluster assignments, restricted to traits shared between the two approaches (n = 162), demonstrated minimal agreement across clusters (maximum adjusted Rand index = 0.0199 at k = 10). Overall, these results indicated that the two clustering approaches organised traits according to largely orthogonal structures, with the limited correspondence observed likely reflecting differences in statistical power rather than shared underlying biology. Across the 19 phenotypes observed in all biobanks, colocalisation analyses implicated 399 unique proteins. Cross-biobank similarity of implicated protein sets varied substantially by phenotype. Six phenotypes exhibited high concordance, with average pairwise Jaccard similarity indices ≥ 0.5: total cholesterol (∼0.71), hematocrit (∼0.64), HDL cholesterol (∼0.62), hemoglobin (∼0.62), red blood cell count (∼0.60), and LDL cholesterol (∼0.58). These phenotypes were characterised by large protein sets with substantial and consistent overlap across biobanks. For example, the three-way intersection for total cholesterol included canonical lipid-related proteins such as APOA1, APOB, APOE, and the regulatory protein ANGPTL3, while proteins including ACE, ADAM23, ENG, IGF1R, INSR, LIFR were shared across lipid and haematological phenotypes. In contrast, anthropometric phenotypes, including height, body mass index, and body weight, showed minimal overlap, with average Jaccard similarity indices below 0.1, indicating substantial heterogeneity in colocalised signals across biobanks.

**Figure 3.**
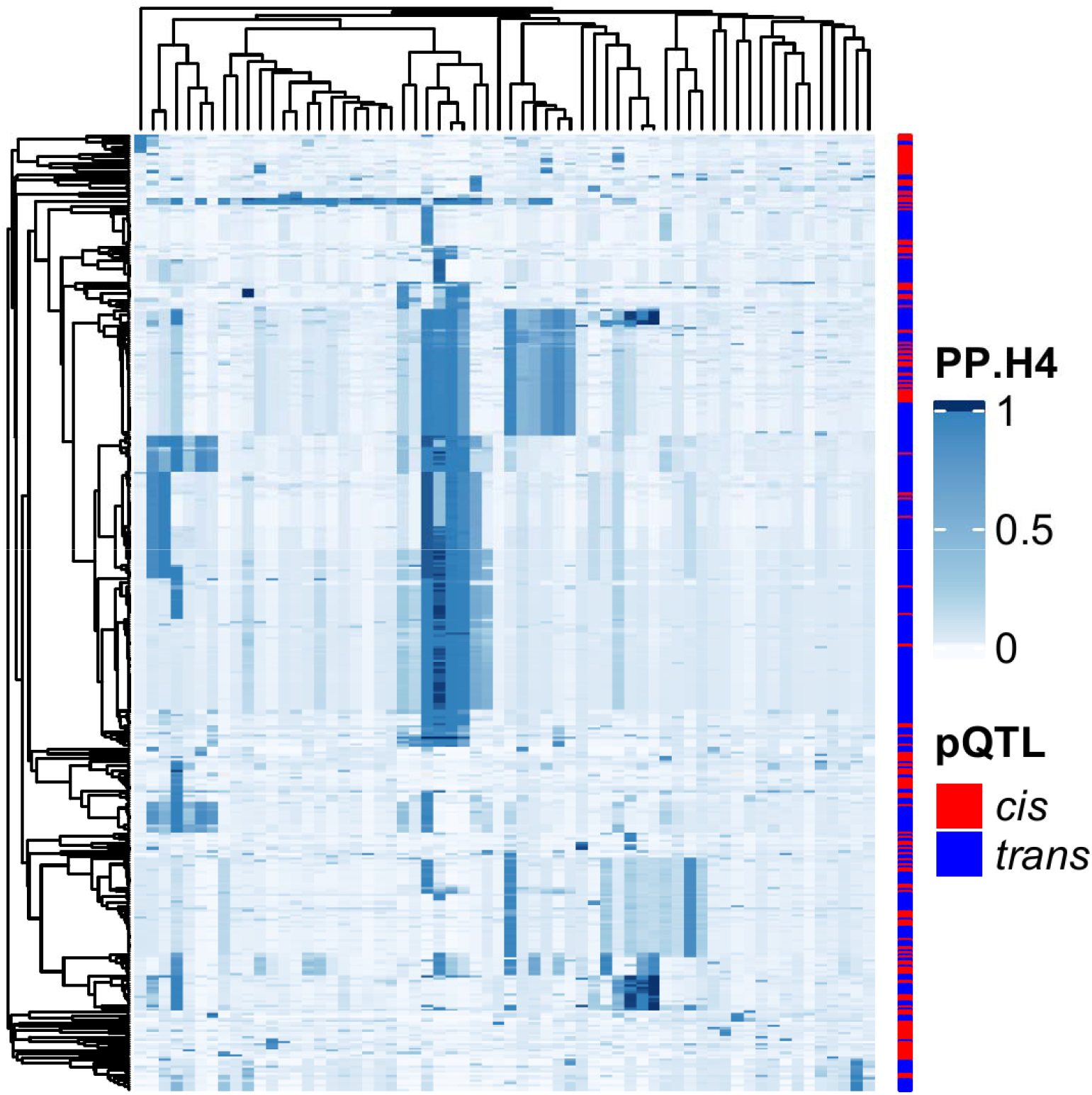
Heatmap of protein-disease colocalisations. Each cell displays the posterior probability of colocalisation (PP.H4), derived from either approximate Bayes factor or *coloc*.*susie*, representing the probability that a protein pQTL and a disease share a causal variant. The heatmap summarises PP.H4 values for 423 proteins (rows) and 62 diseases (columns). Hierarchical clustering using the average-linkage method was applied to group proteins and phenotypes with similar colocalisation profiles. Dendrograms reflect clustering structure.

**Figure 4.**
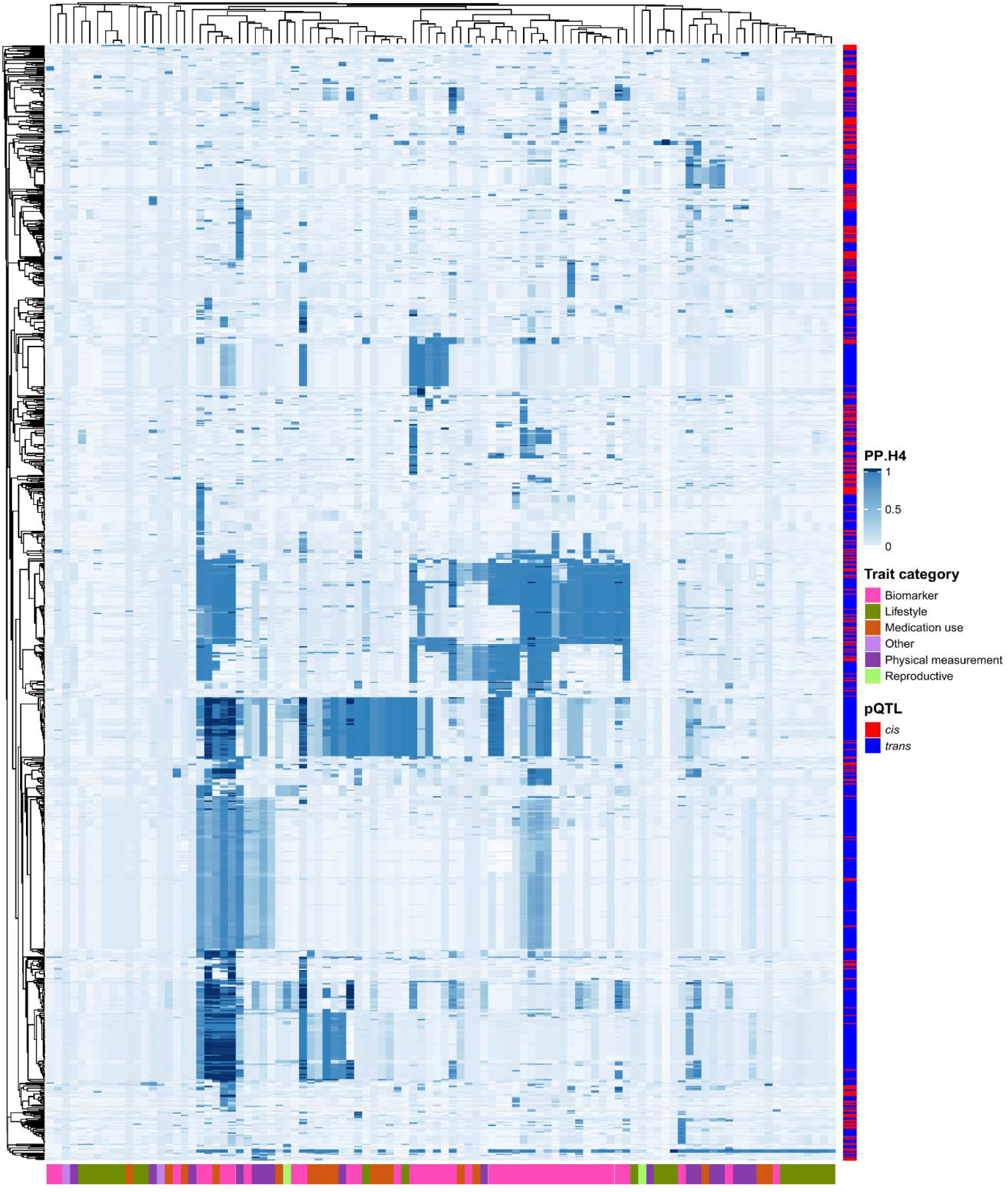
Heatmap of protein-phenotype colocalisations, including medication use. Each cell displays the posterior probability of colocalisation (PP.H4), derived from either approximate Bayes factor or *coloc*.*susie* method, representing the probability that a protein pQTL and a phenotype share a causal variant. The heatmap summarises PP.H4 values for 1,003 proteins (rows) and 100 phenotypes (columns). Hierarchical clustering using the average-linkage method was applied to group proteins and phenotypes with similar colocalisation profiles. Dendrograms reflect clustering structure.

We performed Wald ratio tests and generalised summary-based Mendelian randomisation (GSMR) analyses, restricting genetic instruments to *cis*-pQTLs to minimise horizontal pleiotropy. In total, we identified 1,975 protein-trait pairs (FDR < 0.05), involving 483 proteins and 187 traits. Notably, GSMR identified 21 protein-trait associations that were not detected using single-variant Wald ratio analyses. Of these, seven associations involved the serine protease hepatocyte growth factor activator (HGFAC) and multiple biomarker and anthropometric traits. No significant associations for HGFAC were observed in the Wald ratio analyses, indicating that these findings were driven by the inclusion of multiple independent cis-pQTL instruments in the GSMR framework. Restricting analyses to associations supported by both colocalisation and Mendelian randomisation yielded 645 protein-trait pairs, comprising 222 proteins and 146 traits. Of these, 122 pairs, involving 60 proteins and 48 traits, were related to disease outcomes **(Figure 5)**. While ALDH2 emerged as the most pleiotropic protein, exhibiting associations across a wide range of diseases, we highlight an association between apolipoprotein F (APOF) and hepatic cancer that, to our knowledge, has not been previously identified through genetic or proteome-wide association approaches. Our results indicate that higher genetically predicted levels of APOF are associated with a reduced risk of hepatic cancer (OR_Wald_ = 0.79; OR_GSMR_ =0.78), with strong evidence of colocalisation (PP.H4 = 0.99). The direction of effect is concordant with prior tissue-based and proteomics study reporting down-regulation of APOF in hepatocellular carcinoma and poorer prognosis associated with low APOF expression.^28^

**Figure 5.**
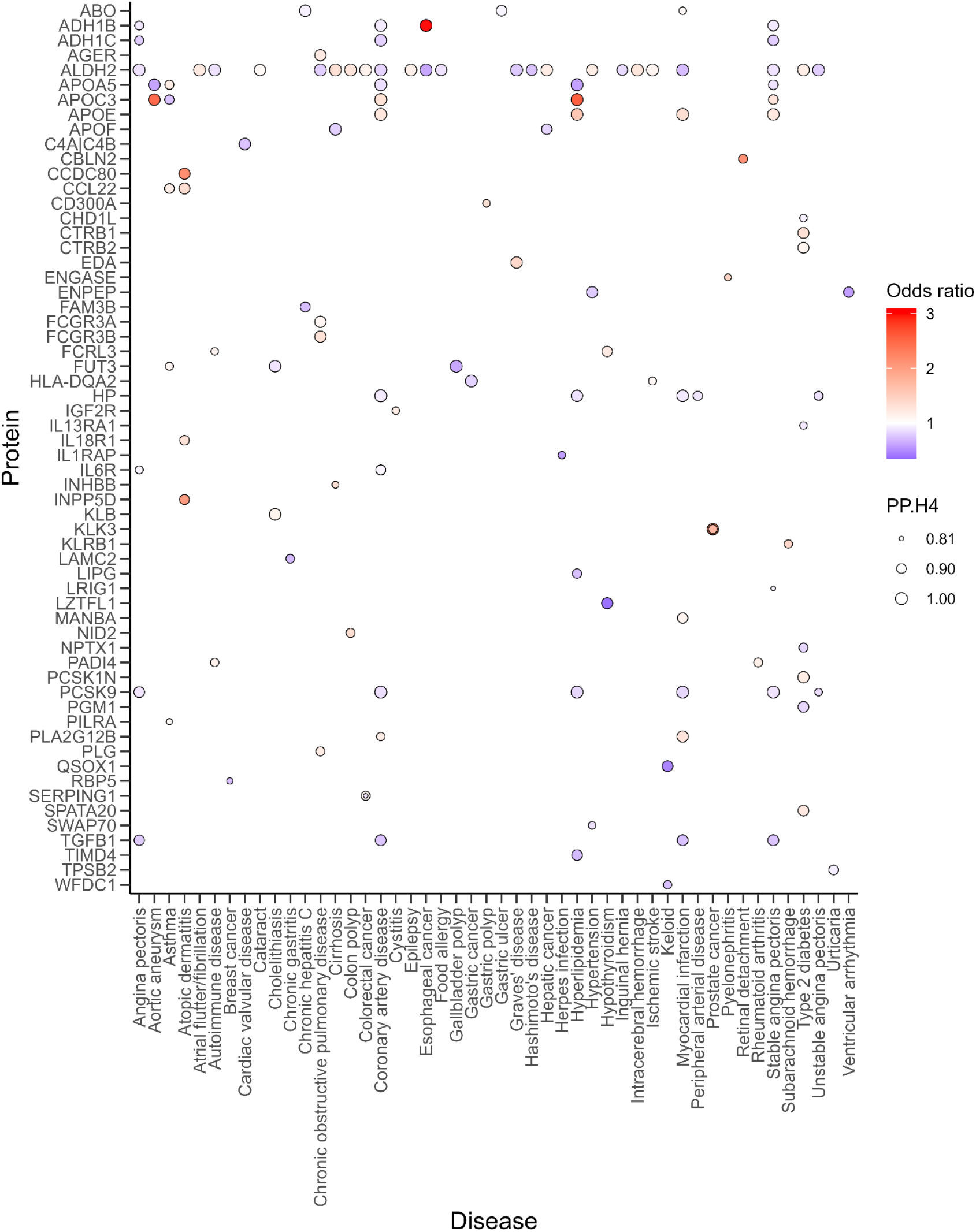
Protein-disease associations supported by both Mendelian randomisation and colocalisation analyses. Rows correspond to proteins and columns to disease outcomes. Circle colour indicates the direction and magnitude of the estimated effect size, with blue representing protective effects and red representing increased risk. Circle size is proportional to the posterior probability of colocalisation (PP.H4), with larger circles indicating stronger evidence for a shared causal variant between protein and disease.

## Discussion

The current study used an integrative framework to systematically characterise the genetic determinants of circulating protein levels in an East Asian population, addressing a major gap in global proteogenomic resources. Approximately one-third of all analysed proteins harboured at least one pQTL and one-sixth had a *cis*-pQTL, largely consistent with proportions reported in European cohorts.^9,19^ Importantly, most lead pQTLs were associated across external cohorts and proteomic platforms, supporting their robustness and validity. This degree of cross-platform and cross-cohort replicability parallels observations from multi-platform comparisons in previous proteogenomic studies.^20-22^ Therefore, we anticipate this resource to facilitate data integration across multiple cohorts^29^ and support the development of new analytical tools for proteomic data.^30^

Although most *cis*-pQTLs in our study showed proximal effects on their encoded proteins, we also identified coherent regulatory networks in which *cis*-acting variants influenced distal *trans*-protein effects. Similar *trans*-mediated networks have been observed in the past across large-scale proteogenomic studies.^19^ We identified several prominent hubs forming *trans*-driven networks, involving apolipoproteins (APOE, APOC1 and APOB), complement system components (CFH/CFHR family members, C1R/C1S, C4A/C4B, and collectins), coagulation-related proteins (F5, HRG, KNG1, and VTN) and circulating carrier proteins. The gene-based analyses in UKB-PPP suggest that many more genotype-protein hubs involving rare variants exist^31^, many of which are not detectable through standard GWAS frameworks.

From a translational perspective, our proteome-phenome-wide atlas facilitates prioritisation of candidate biomarkers and targets by integrating evidence of genetic regulation, colocalisation with phenotypes and diseases, and putative causal inference via Mendelian randomisation. Phenome-wide evaluation of protein associations revealed sparse yet structured landscape, with relationships predominantly driven by *trans*-pQTLs. Although increasing number of studies have used *trans*-pQTLs to infer causality,^13,32,34^ the usefulness and robustness of this approach remain to be systematically assessed. There is emerging evidence that, unlike *cis*-pQTLs, *trans*-pQTLs are largely tissue-specific;^35,36^ however, confirming this pattern and elucidating how *trans*-pQTL mediated regulation operates and interacts in biologically coherent networks will require further studies across a broader and more diverse range of tissues. Our proteome-phenome-wide analyses identified several well-established clusters, including a prominent cardiometabolic cluster, for which our study had the greatest statistical power. While the PheWAS and Mendelian randomisation analyses identified protein-trait associations across nearly every biological domain, the colocalisation analyses indicated that only a subset of these associations have shared causal variants. Although discrepancies could arise from differences in underlying biology or from technical artefacts such as LD mismatch, the most plausible explanation is limited statistical power in the outcome GW AS. Consistent with this, most shared signals were observed for quantitative phenotypes, such as lipid levels, blood pressure, and anthropometric measures, whereas shared signals for binary outcomes were markedly depleted. These observations underscore the need for more highly powered GWAS in non-European populations to enable a more complete delineation of the genomic architecture underlying protein-trait associations.

We have highlighted an association between APOF and hepatic cancer. A biologically plausible explanation for the observed inverse association between APOF and hepatic cancer risk lies in APOF’s central role in hepatic lipid metabolism. APOF is predominantly expressed in the liver and regulates lipoprotein remodelling and cholesterol transport, processes that are tightly linked to hepatic metabolic homeostasis. Dysregulation of lipid handling and chronic metabolic inflammation are key drivers of hepatocarcinogenesis, and reduced APOF expression may exacerbate lipotoxicity, oxidative stress, and inflammatory signalling within hepatocytes. Consistent with this hypothesis, prior tissue-based study has reported down-regulation of APOF in hepatocellular carcinoma and poorer prognosis associated with low APOF expression.^28^ Our Mendelian randomisation results extend these observations by providing genetic evidence that lifelong higher APOF levels are associated with reduced hepatic cancer risk, supporting a potential protective role for APOF in liver carcinogenesis.

Our study has several strengths. We extend the current proteogenomic landscape to a non-European population with the largest sample size and the greatest number of proteins assayed to date. Consistent with a recent multi-ancestry proteome-phenome MR results^14^ we found proteins that were instrumentable only in East Asians and that such ancestry-specific instruments contributed to hundreds of protein-trait associations that could not have been detected using European data alone. Therefore, the current study adds to a limited number of available East Asian proteogenomic studies^25,37,38^ and provides a resource that can be leveraged to identify potentially novel protein-phenotype relationships. The use of two independent proteomic platforms in the same individuals enabled cross-platform validation of associations, increasing confidence in the robustness of our findings. Integrating genetic and proteomic data with phenotypic information from multiple cohorts allowed us to discover insights that would not have been feasible in smaller or single-cohort studies. Nonetheless, several limitations warrant consideration. A proportion of our pQTLs did not replicate in European cohorts, indicating that some pQTLs may be driven by protein-altering variants (PAVs) that influence epitope binding or they could be population-specific. Epitope effects have been quantified to account for approximately 12% of *cis*-pQTLs and may reflect missense variants directly affecting reagent binding rather than true protein abundance.^39^ Similar analyses in cross-ancestry comparisons indicate that the prevalence and impact of such artefacts differ across populations due to allele-frequency differences in missense variants.^40^ The SomaScan assay, while broad, does not capture the full proteome and exhibits variable sensitivity, which may bias pQTL discovery. Moreover, Mendelian randomisation and colocalisation analyses rest on assumptions that may be violated in the presence of unmeasured confounding, horizontal pleiotropy, or cross-population differences in LD structure. Therefore, even with stringent quality control, some associations may require functional validation.

In conclusion, the present East Asian proteogenomic atlas substantially expands the known landscape of pQTLs in an underrepresented ancestry and provides a valuable reference for interpreting protein-trait relationships across populations. When viewed alongside large proteomic resources from European, Icelandic, Japanese, Chinese, and Arctic cohorts, our data reinforce the importance of ancestry-inclusive proteomics for biomarker development, drug target prioritisation, and mechanistic insight. The atlas generated here will enable future studies to delineate causal protein pathways, refine therapeutic hypotheses, and improve the calibration and interpretability of proteomic platforms across ancestries. Further expansions into rare-variant analyses, variance QTLs, multi-omics integration, and better-powered non-European GWAS will be critical to fully resolve the proteogenomic architecture that underlies human disease.

## Acknowledgements

The chief acknowledgment is to the study participants, the members of the survey teams in each of the 10 regional centres, and the project development and management teams based at Beijing, Oxford and the 10 regional centres. We thank Sarah Clark, Martin Radley, and Mike Hill at CTSU, Oxford, for assisting with the planning and organisation of the proteomics measurements.

The CKB baseline survey and the first re-survey were supported by the Kadoorie Charitable Foundation in Hong Kong. The long-term follow-up and subsequent resurveys have been supported by Wellcome grants to Oxford University (212946/Z/18/Z, 202922/Z/16/Z, 104085/Z/14/Z, 088158/Z/09/Z) and grants from the National Natural Science Foundation of China (82192901, 82192904, 82192900) and from the National Key Research and Development Program of China (2016YFC0900500).The UK Medical Research Council (MC_UU_00017/l, MC_UU_12026/2, MC_U137686851), Cancer Research UK (C16077/A29186, C500/A16896) and the British Heart Foundation (CH/1996001/9454), provide core funding to the Clinical Trial Service Unit and Epidemiological Studies Unit at Oxford University for the project. The proteomic assays were supported by BHF (18/23/33512), Novo Nordisk, and Olink. DNA extraction and genotyping were supported by GlaxoSmithKline and the UK Medical Research Council (MC-PC-13049, MC-PC-14135). Computation used the Oxford Biomedical Research Computing (BMRC) facility, a joint development between the Wellcome Centre for Human Genetics and the Big Data Institute supported by Health Data Research UK and the NIHR Oxford Biomedical Research Centre; the views expressed are those of the authors and not necessarily those of the NHS, the NIHR, or the Department of Health.

## Methods

### Study population

Details of the study design of China Kadoorie Biobank (CKB) have been previously reported elsewhere.^41,42^ In summary, during 2004 and 2008, 512,726 Chinese adults aged 30-79 years were recruited from the general population in 5 geographically defined urban and 5 rural areas of China. Questionnaire data, physical measurements, and blood samples were collected by trained health workers at local assessment centres. Additional data were collected at three follow-up surveys, involving approximately 5% randomly selected surviving participants. The current study included 1976 incident cases of IHD and 2,001 subcohort participants. Cases of IHD were defined based on International Classification of Diseases, Tenth Revision (ICD-10) codes 120-125. These cases were accrued over a 12year follow-up before 1 January 2019 and comprised a random sample that had genome-wide data, no history of cardiovascular disease, and no statin use at baseline. Subcohort participants were randomly selected from a population subset of 69,353 genetically unrelated participants that had genome-wide data and no history of IHD or stroke and no statin use at baseline **(Supplementary Table 1)**.

### Proteomic profiling

Retrieved plasma samples for 3,977 participants were aliquoted into 96-well plates (11 wells allocated for external control samples, including 5 calibrator, 3 QC, and 3 buffer samples) and sent to SomaLogic for profiling using SomaScan assay v4.1. The 120 ul aliquots were split into two and run in duplicate. All samples were barrel randomized before the assay. Two aliquots had too little volume to run in the first instance, and three more had too little sample volume to be run in duplicate. Only one measure for each duplicated reagent was kept in our analysis. To achieve optimal detection of protein targets, three serial dilutions (0.05%, 0.5%, and 20%) were conducted in the SomaScan assay. The raw assay data were standardised using external control samples to mitigate variability arising from microarrays and variations within and across plates by SomaLogic. Utilising external reference, an additional optional step of adaptive normalisation using maximum likelihood (ANML) was performed to control for intersample variability.^22,27^ The final SomaScan data were supplied in both ANML and non-ANML versions in relative fluorescence units (RFU). QC checks were performed by comparing the median of QC samples on each plate to the reference, and a cross-plate QC check measure (pass/flag) was assigned to each aptamer. In SomaScan assay v4.1, measurements for 7,596 aptamers were available, which included 7,289 human protein targets, 307 non-human aptamers, of which the vast majority were mouse-related proteins (n = 236), and 71 aptamers targeting non-proteins such as spuriomer, non-biotin and hybridisation control elusion. Overall, the ANML-normalised data yielded stronger variant-protein associations **(Supplementary Figure 1a)** and identified a greater number of significant pQTLs than the non-normalised data **(Supplementary Figure 1b)**. Consequently, we used the ANML dataset for the main analyses (aptamer metadata are provided in **Supplementary Table 2)**.

### Phenotype derivation and sample exclusion

For each version of SomaScan data (non-ANML and ANML), aptamer measurements were log-transformed and then adjusted for age, age^2^, sex, study region, and national principal components using linear regression. Regression residuals were subsequently rank-based inverse normal transformed (RINT) to approximate normality. These RINT-transformed residuals were subjected to principal component analysis, and individuals identified as outliers (>5 SD from the mean in any of the first 10 principal components) were removed. This procedure identified 8 outliers in the non-ANML dataset and 12 outliers in the ANML dataset. After excluding these individuals, final protein phenotypes for 3,969 non-ANML and 3,965 ANML participants were derived by repeating the linear regression on log-transformed protein levels using the same covariates, followed by extraction and RINT transformation of the residuals.

### Genotyping and imputation

Genotyping of CKB participants has been previously described in Walters *et al*^43^ In brief, genotyping of samples was performed on a custom Affymetrix Axiom array designed by CKB and Affymetrix, with input from Beijing Genomics Institute (BGI). The array included a GWAS scaffold optimized for imputation performance in Chinese populations, GWAS catalogue hits, putative loss-of-function variants, and other modules and provided genome-wide coverage for both common and low-frequency variants. Genotypes of 531,565 variants passing QC in all 100,706 genotyped samples were converted to genome build 38 using CrossMap (vO.6.144)^44^ and were checked for consistency by reversing the process (“liftUnder”). Variants not mapped, mapped to different chromosomes, or not mapped back to the same locations after liftUnder, were excluded. The remaining 531,542 variants were prephased using SHAPEIT v4.2 (SHAPEIT v2.904 for chromosome X) and imputed with Westlake BioBank for Chinese (WBBC) and *Trans*-Omics for Precision Medicine (TOPMED) reference panels. The two sets of imputed data were then merged, for each variant retaining the imputed genotypes with the higher imputation INFO score. SNPs with call rates <98%, Hardy-Weinberg equilibrium P-value < 10^-7^ or minor allele frequency (MAF) =0 were excluded.

### Genome-wide association study

The GWAS for 7,596 SomaScan aptamers in ∼4,000 CKB participants were performed using *REGENIE*^*45*^ (v3.2.5.2) involving both ANML and non-ANML datasets, and assuming an additive inheritance model.We adjusted for array, age, age^2^, sex, study region, and 11 national principal components. Approximately 12 million SNPs with imputation score >0.3 and minor allele count >20 were included in analyses. The genomic region boundaries including genome-wide significant pQTLs (P<5×10^-8^) were defined by LD-based clumping in *PLINK*^*46*^ (initial window +/-5Mbp, P<0.05, LD r^2^>0.05) using an internal LD reference of 72,000 unrelated CKB participants with imputed genotype probabilities converted to “best-guess” genotypes. For the HLA region (chr6:21744977-39074734) an initial window was set to +/-20Mbp. The overlapping genomic boundaries were merged and extended by 10kb, and the variant with the lowest P-value was identified as the sentinel pQTL. pQTLs that were within 500kb on either side of the protein-encoding gene were defined as *cis*-pQTLs and those lying outside were defined as *trans-* pQTLs. Non-human proteins were defined neither as *cis* or *trans-*pQTLs. The lead pQTLs identified in this study were replicated using CKB OLINK pQTLs,^25^ UKB-PPP OLINK pQTLs,^19^ and deCODE SomaScan v4.0 pQTLs.^9^ The exact conditional analysis was used to identify secondary pQTL associations. Using *cis*-pQTL instruments, we identified instances in which the same genetic variant exerted both *cis* and *trans* effects, with significance at P < 6.58 x I0^-13^. These relationships were subsequently used to construct a protein-protein regulatory network.

### Fine-mapping and functional annotation of pQTLs

Fine-mapping within clumped and merged pQTL loci used SuSIE (v0.14.2) as implemented in R package susieR^47^ to obtain CSs of likely causal variants with a total PIP >0.95, using individual level data for transformed protein levels and genotype dosages. The maximum number of credible sets (L) was determined by application of the SuSIE_auto function (modified to iteratively increase *L* by 1 rather than doubling) after which the results were filtered to remove low-quality CSs failing to reach the specified tolerance (0.001) or those which did not include a variant reaching the specified significance threshold (6.58 x 10^13^). The variant with the largest PIP in each independent CS was classed as its lead variant. Variant annotation of fine-mapped pQTLs was conducted according to Ensembl VEP 107.^48^ We selected the most severe annotation for all variants across all available transcripts using *BCFtools*^*49*^ command ‘split-vep -s worst’. Variants were categorised according to their functional impact and location with respect to coding and transcript sequences, and according to whether they affected the gene encoding the target (i.e. assayed) protein, a gene for a different protein within the *cis* region, or at a *trans*-pQTL. Variants that had functional annotation as splice acceptor/donor, start lost, stop gained/lost, frameshift, missense, in-frame insertion/deletion, and protein altering variant were considered to be PAVs.

### PheWAS, colocalisation and Mendelian randomisation

We performed PheWAS lookups of lead pQTL SNPs (P < 6.58 x 10^-13^) or their proxies (r^2^ > 0.8 within an LD-clumped window) across 225 traits obtained from BioBank Japan (BBJ), the Korean Genetic Epidemiology Study (K0GES) or Taiwan biobank (TWB). All datasets were aligned to the GRCh37 genome build. Colocalisation analyses were carried out for all SNPs within the LD-clumped window using both the approximate Bayes factor and SuSiE approaches implemented in *coloc* (v5.2.1)^50^, with the internal CKB LD reference. For each protein-phenotype pair, we considered evidence of colocalisation when the posterior probability of a shared causal variant (PP.H4) exceeded 0.8 and the ratio PP.H4 / PP.H3 exceeded 5. Sensitivity analyses were conducted to confirm that colocalisation signals were robust to variation in prior probabilities. Using the lead *cis*-pQTLs or their proxies (r^2^ > 0.8 within an LD-clumped window, we performed Wald ratio tests to estimate the effect of each circulating protein on outcome traits using the TwoSampleMR (v0.5.5).^51^ All *cis*-pQTLs or their proxies had F-statistics > 10. Steiger directionality tests were applied to assess whether each SNP explained more variance in the exposure than in the outcome. Where possible, we also applied generalised summary-data-based mendelian randomisation v2 (GSMR2)^52^ to conduct multi-variant Mendelian randomisation while accounting for LD among instruments. Protein-trait associations were considered significant at a false discovery rate (FDR) < 0.05. Hierarchical clustering was used to group proteins and traits with similar association profiles. Cophenetic correlation analysis indicated that the “average” linkage method was most appropriate across all analyses. For downstream heatmap visualisation using the ComplexHeatmap package in R,^53^ we selected results from coloc.abf or coloc.susie (whichever yielded the higher PP.H4) and from either the Wald ratio or GSMR (whichever yielded the lower FDR). We compared our findings against previously published phenome-proteome-wide datasets to determine which of our protein-trait associations represented potentially novel signals. For each phenotype observed in all three biobanks (BBJ, K0GES and TWB), we constructed biobank-specific sets of implicated proteins based on colocalisation results. To compare clusters derived from PheWAS and colocalisation analyses, agreement between clusters was quantified using the adjusted Rand index (ARI) across a range of cluster numbers (k = 2-30). ARI was computed after restricting both dendrograms to their intersection of traits to ensure comparability, and the maximum ARI observed across all values of k was reported. Cross-biobank similarity was quantified using the Jaccard similarity index, defined for each biobank pair as the size of the intersection divided by the size of the union of the corresponding protein sets. For each phenotype, Jaccard indices were calculated for all three biobank pairs (BBJ-K0GES, BBJ-TWB and K0GES-TWB), and the average of these three values was used as a summary measure of cross-biobank concordance.

## Authors’ contributions

AP analysed the data. AP, BW, IYM, RW, ZC drafted the manuscript. AP, IYM, LL, ZC and RGW contributed to the conception of this paper, interpretation of the results and the revision of manuscript. RGW, LL and ZC designed the study. KL, MY, YG, CM, JL, CY, DA, DSV, LL, ZC, IYM, and RGW, contributed to data acquisition. All authors critically reviewed the manuscript and approved the final submission.

## Data availability statement

Summary statistics reported in this paper have been deposited to GWAS Catalog (accession codes GCST90718748-GCST90726343), and will also become available for download on the CKB PheWeb browser (pheweb.ckbiobank.org), and have been deposited in the Genome Variation Map (GVM) at the National Genomics Data Center, China National Center for Bioinformation under accession number GVP000045. CKB non-genetic data are available and updated periodically for access by bona fide researchers. Details of the CKB Data Sharing Policy, data release schedules and data request application procedures are available at www.ckbiobank.org. Queries about data access should be addressed to. Access to individual participant genetic data is currently constrained by China’s Administrative Regulations on Human Genetic Resources, for which collaboration with CKB researchers is generally required, and which may be subject to separate regulatory approvals in China if it involves substantial sharing of unpublished data. Summary statistics for East-Asian traits used in this study were downloaded from https://pheweb.jp/downloads (BBJ),^54^ https://koges.leelabsg.org/ (K0GES).^55^ Summary statistics for Taiwan Biobank (TWB) were obtained from the GWAS catalog.^56^

## Ethics statement

Ethical approval was obtained from the Ethical Review Committee of the Chinese Centre for Disease Control and Prevention (Beijing, China, 005/2004) and the Oxford Tropical Research Ethics Committee, University of Oxford (UK, 025-04), and all participants provided written informed consent.

## Open access statement

This research was funded in whole, or in part, by the Wellcome Trust [212946/Z/18/Z, 202922/Z/16/Z, 104085/Z/14/Z, 088158/Z/09/Z], For the purpose of Open Access, the author has applied a CC-BY public copyright licence to any Author Accepted Manuscript version arising from this submission.

