## Supplementary Figures and Tables for "Genomic atlas of 7,000 plasma proteins and their associations with diseases and traits in East Asian populations": Supplementary Figures.docx

**Supplementary Figure 1. Flowchart summarising the main analytical approaches used in the study and the corresponding key findings at each analytic stage.**

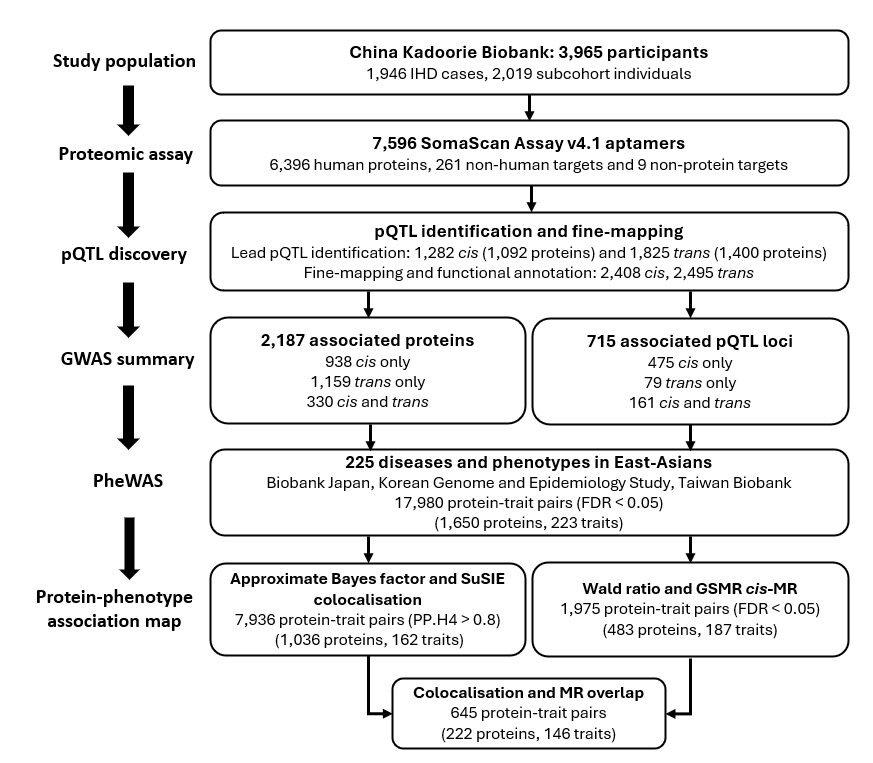

**Supplementary Figure 2. Evaluation of differences in GWAS identified pQTLs between the ANML and non-ANML datasets.** (A) Comparison of lead pQTL (*P* < 6.58 x 10^-13^) associations. (B) Total number of pQTLs identified per SomaScan aptamer.

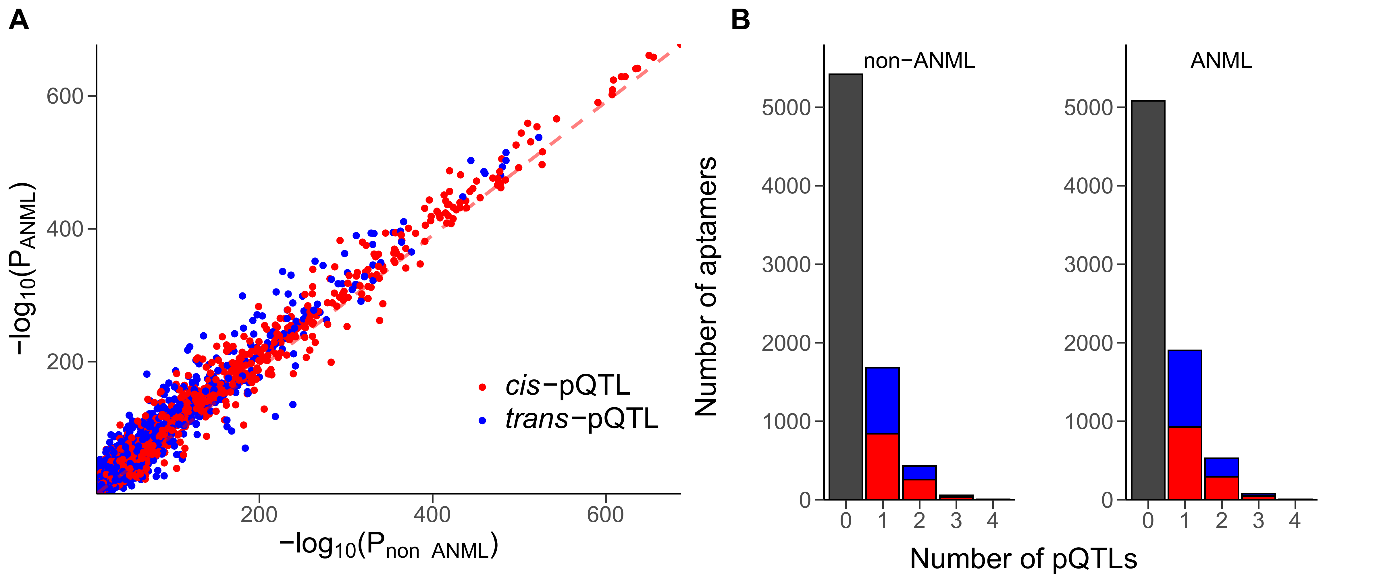

**Supplementary Figure 3. Comparison of lead pQTLs identified using CKB SomaScan with corresponding associations in (A) the UKB-PPP Olink dataset and (B) the CKB Olink dataset.**

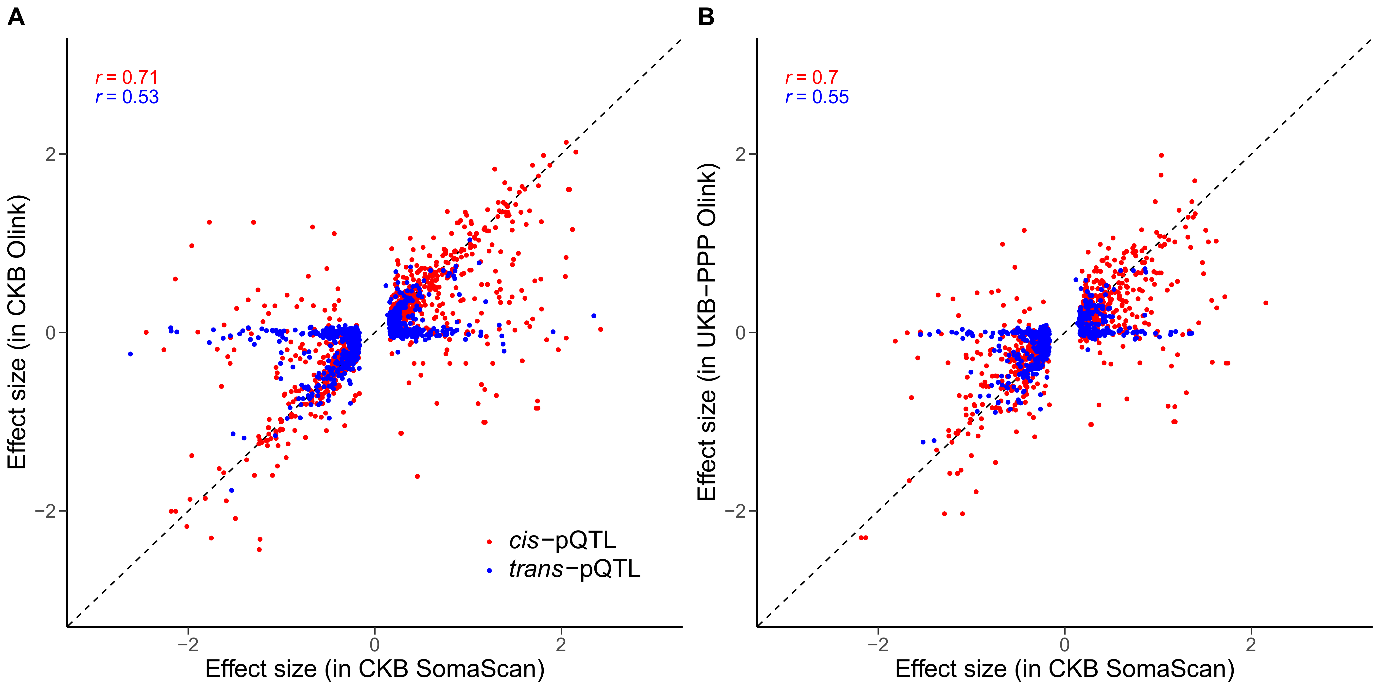

**Supplementary Figure 4. Comparison of effect allele frequencies in CKB with those reported in external East Asian biobanks.**

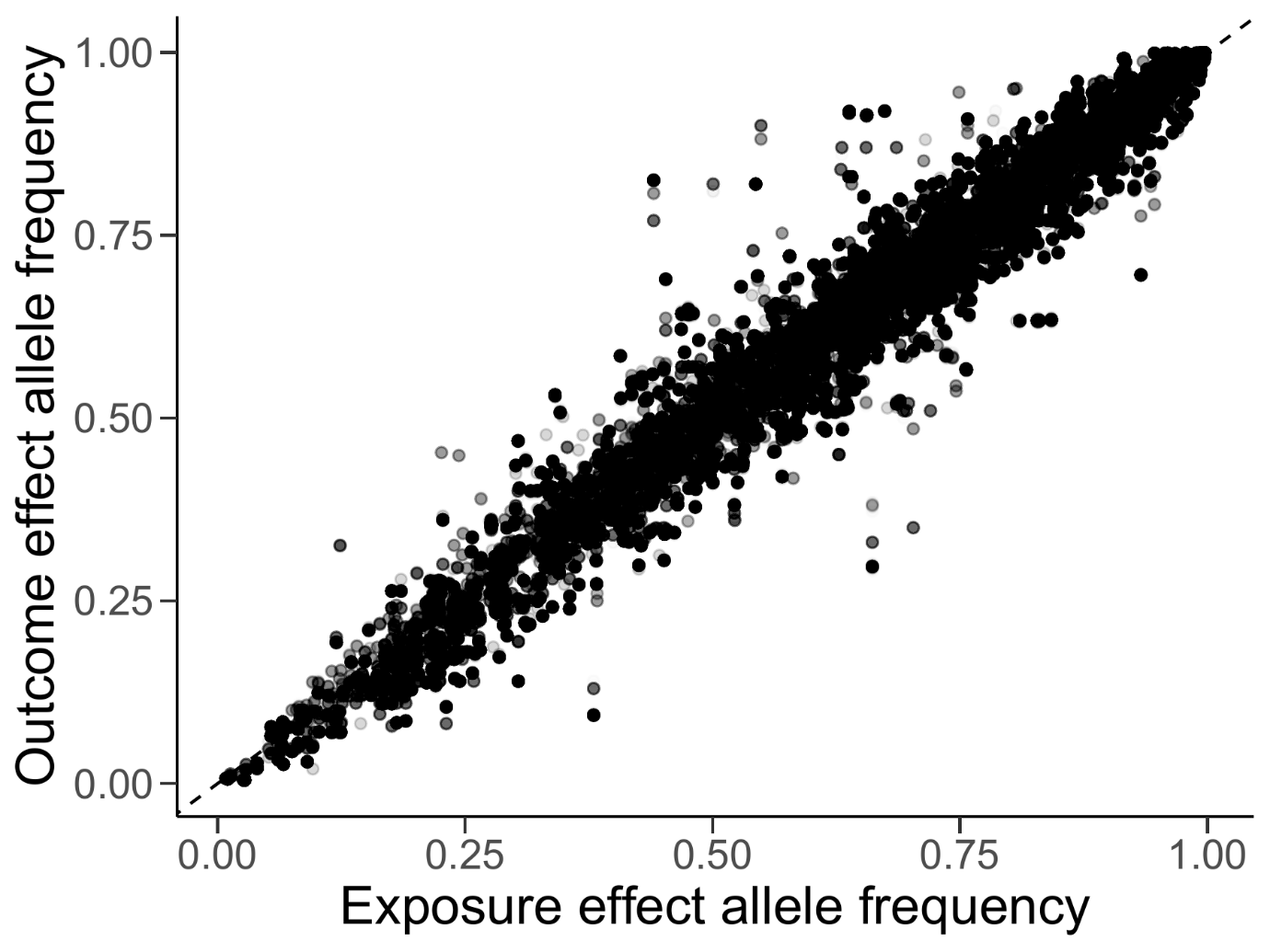

**Supplementary Figure 5. Heatmap of protein-disease PheWAS results, including medication use phenotypes.** Each cell shows the *z*-score for the association between protein levels (rows) and disease or medication use phenotypes (columns). Positive *z*-scores (red) indicate higher protein levels associated with higher phenotype risk or medication use; negative *z*-scores (blue) indicate the opposite direction. Hierarchical clustering using the average-linkage method was applied to group proteins and phenotypes with similar association profiles, and the dendrograms reflect the resulting clustering structure. Trait categories and cis/trans pQTL classifications are shown as side annotations. Trait categories and cis/trans pQTL classifications are shown as side annotations.

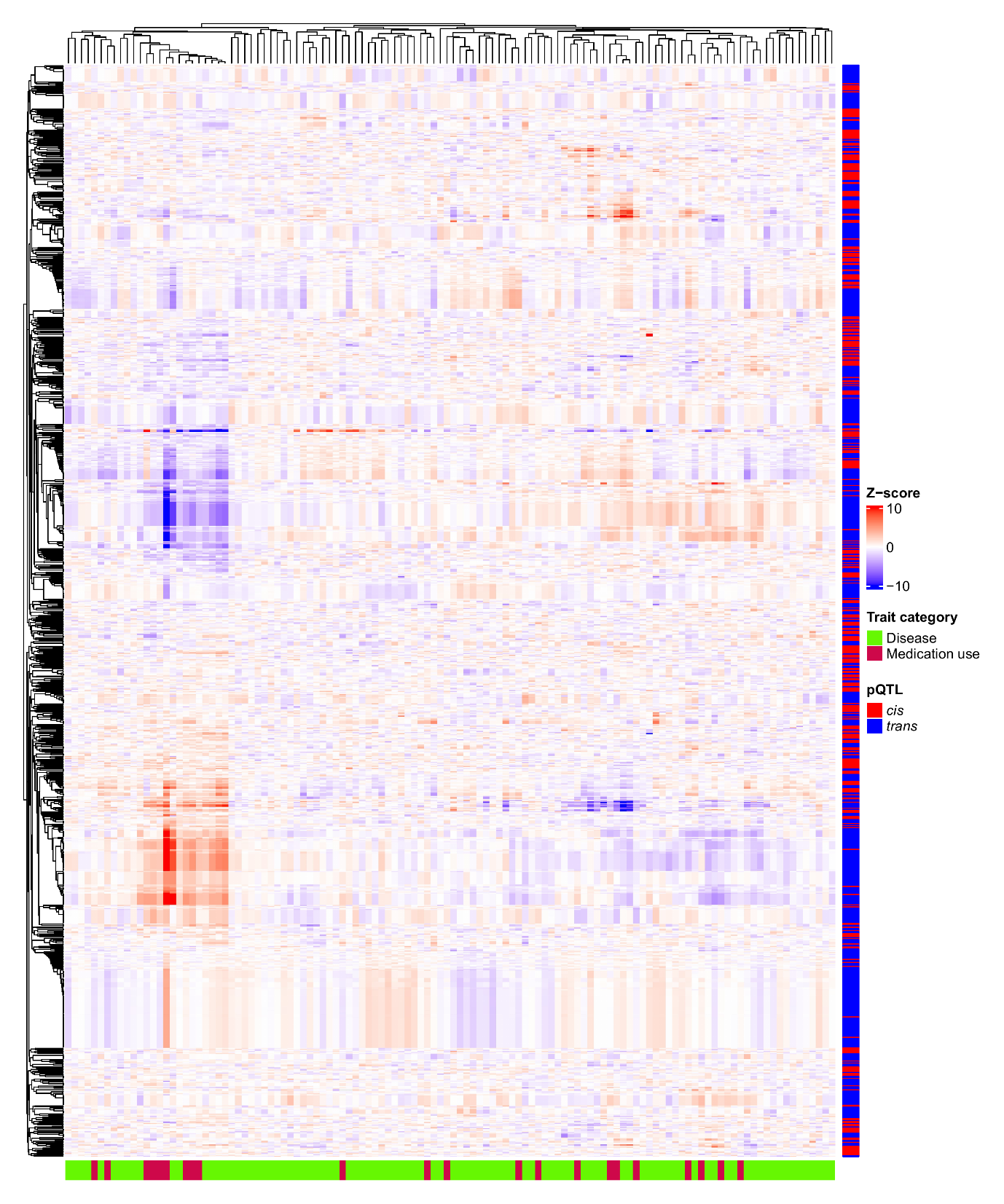

**Supplementary Figure 6. Heatmap of protein-phenotype PheWAS results.** Each cell shows the *z*-score for the association between protein levels (rows) and disease or medication use phenotypes (columns). Positive *z*-scores (red) indicate higher protein levels associated with higher phenotype risk or medication use; negative *z*-scores (blue) indicate the opposite direction. Hierarchical clustering using the average-linkage method was applied to group proteins and phenotypes with similar association profiles, and the dendrograms reflect the resulting clustering structure. Trait categories and cis/trans pQTL classifications are shown as side annotations. Trait categories and cis/trans pQTL classifications are shown as side annotations.
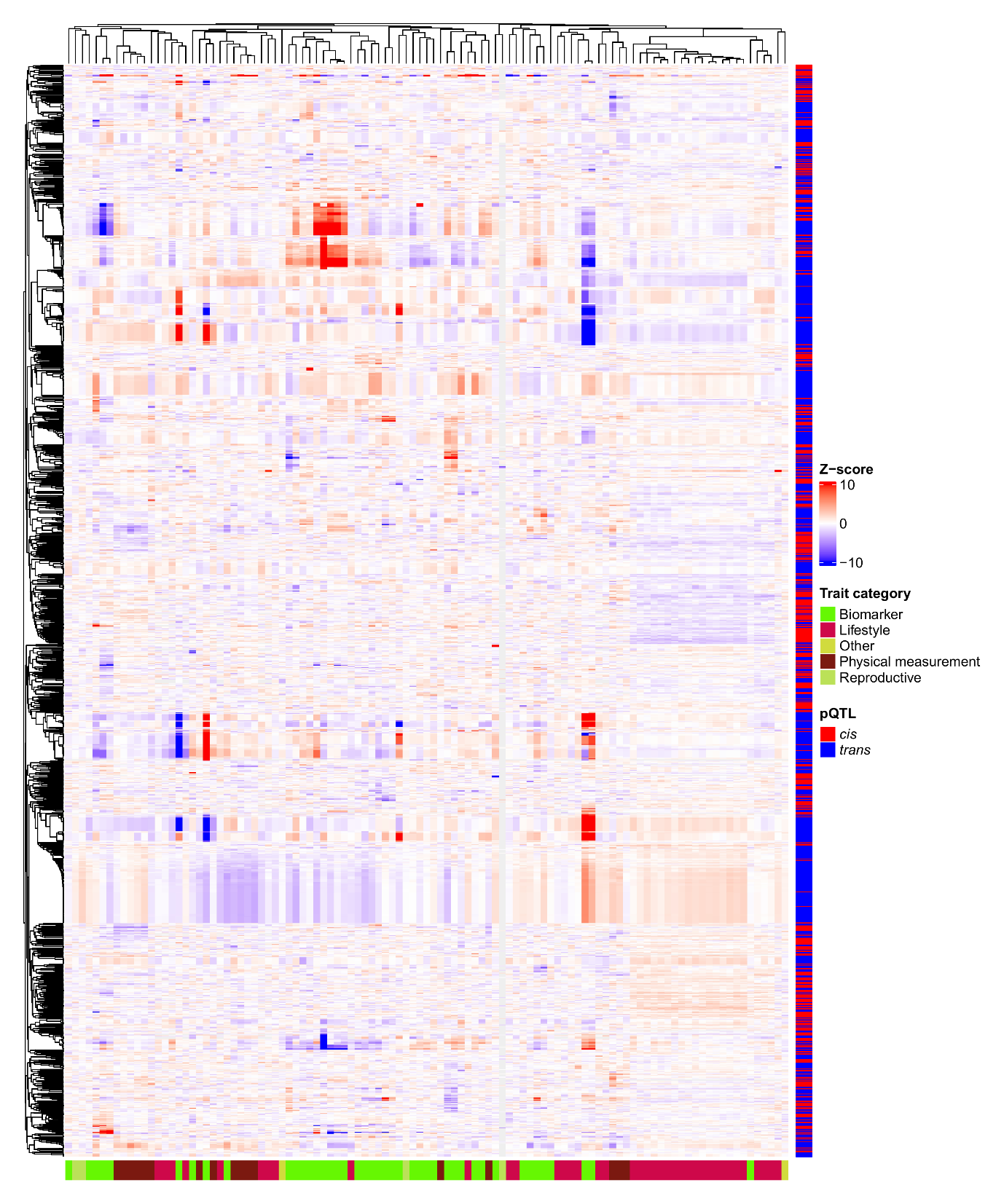

**Supplementary Figure 7. Combined heatmap of protein–disease and protein–phenotype PheWAS associations, including medication-use phenotypes.** Each cell shows the *z*-score for the association between protein levels (rows) and disease or medication use phenotypes (columns). Positive *z*-scores (red) indicate higher protein levels associated with higher phenotype risk or medication use; negative *z*-scores (blue) indicate the opposite direction. Hierarchical clustering using the average-linkage method was applied to group proteins and phenotypes with similar association profiles, and the dendrograms reflect the resulting clustering structure. Trait categories and cis/trans pQTL classifications are shown as side annotations. Trait categories and cis/trans pQTL classifications are shown as side annotations.

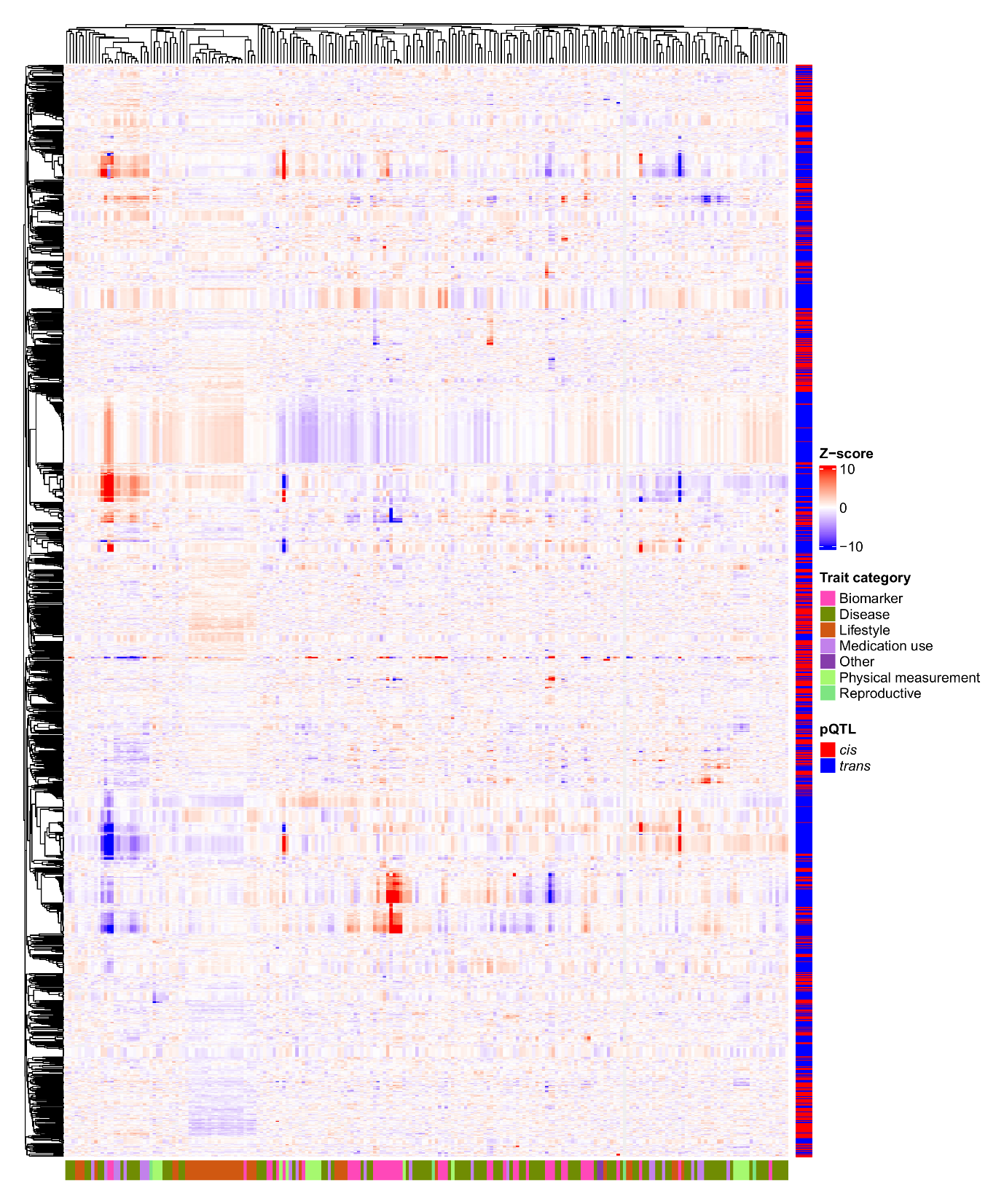

**Supplementary Figure 8. Dendrograms for trait clusters based on PheWAS *z*-scores.** (A) Dendrogram for diseases and medications. (B) Dendrogram for phenotypes. (C) Combined dendrogram including diseases, medications, and phenotypes.

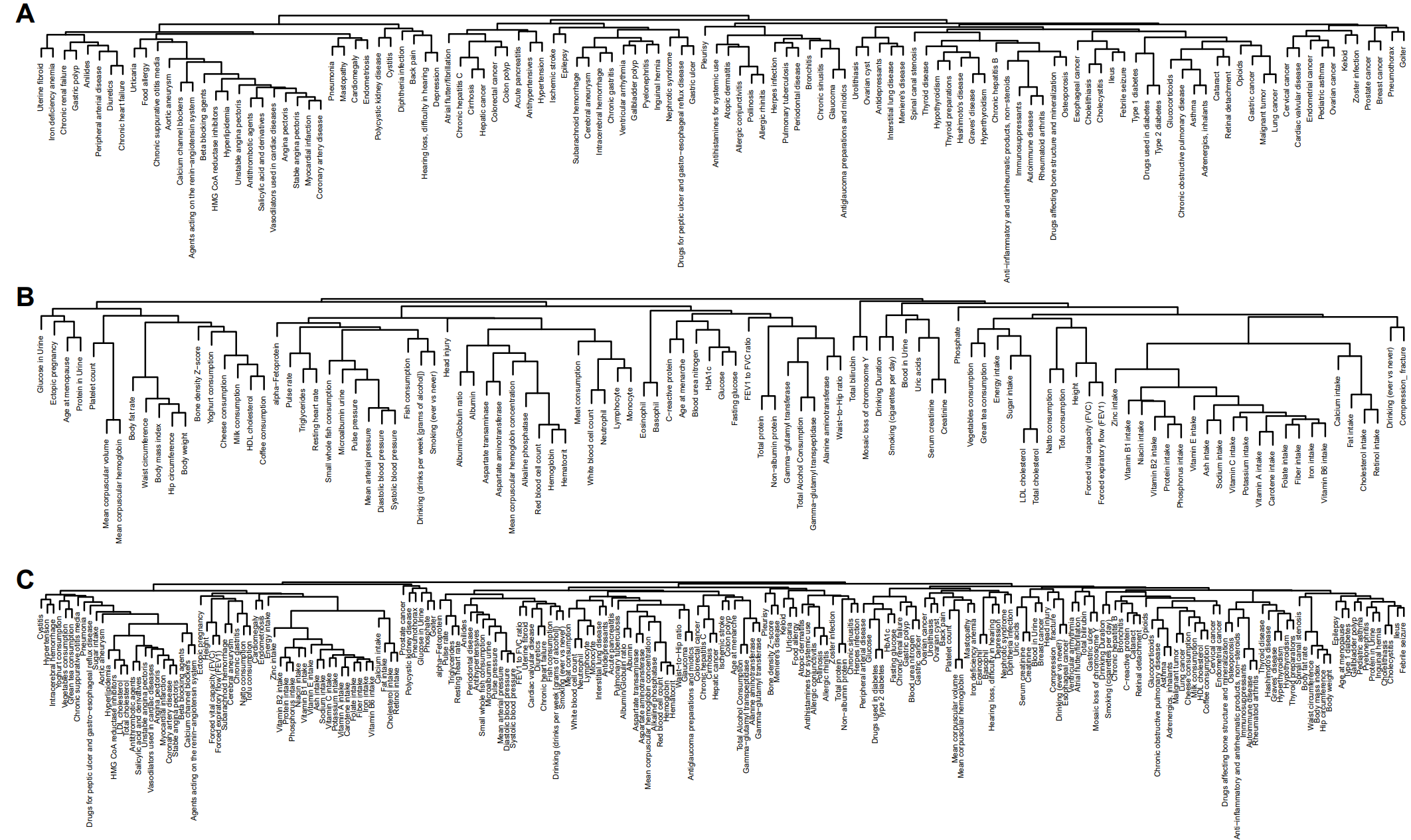

**Supplementary Figure 9. Heatmap of protein-trait colocalisations.** Each cell displays the posterior probability of colocalisation (PP.H4), derived from either approximate Bayes factor or *coloc.susie* method, representing the probability that a protein pQTL and a phenotype share a causal variant. The heatmap summarises PP.H4 values for 1,036 proteins (rows) and 162 phenotypes (columns). Hierarchical clustering using the average-linkage method was applied to group proteins and phenotypes with similar colocalisation profiles. Dendrograms reflect clustering structure.

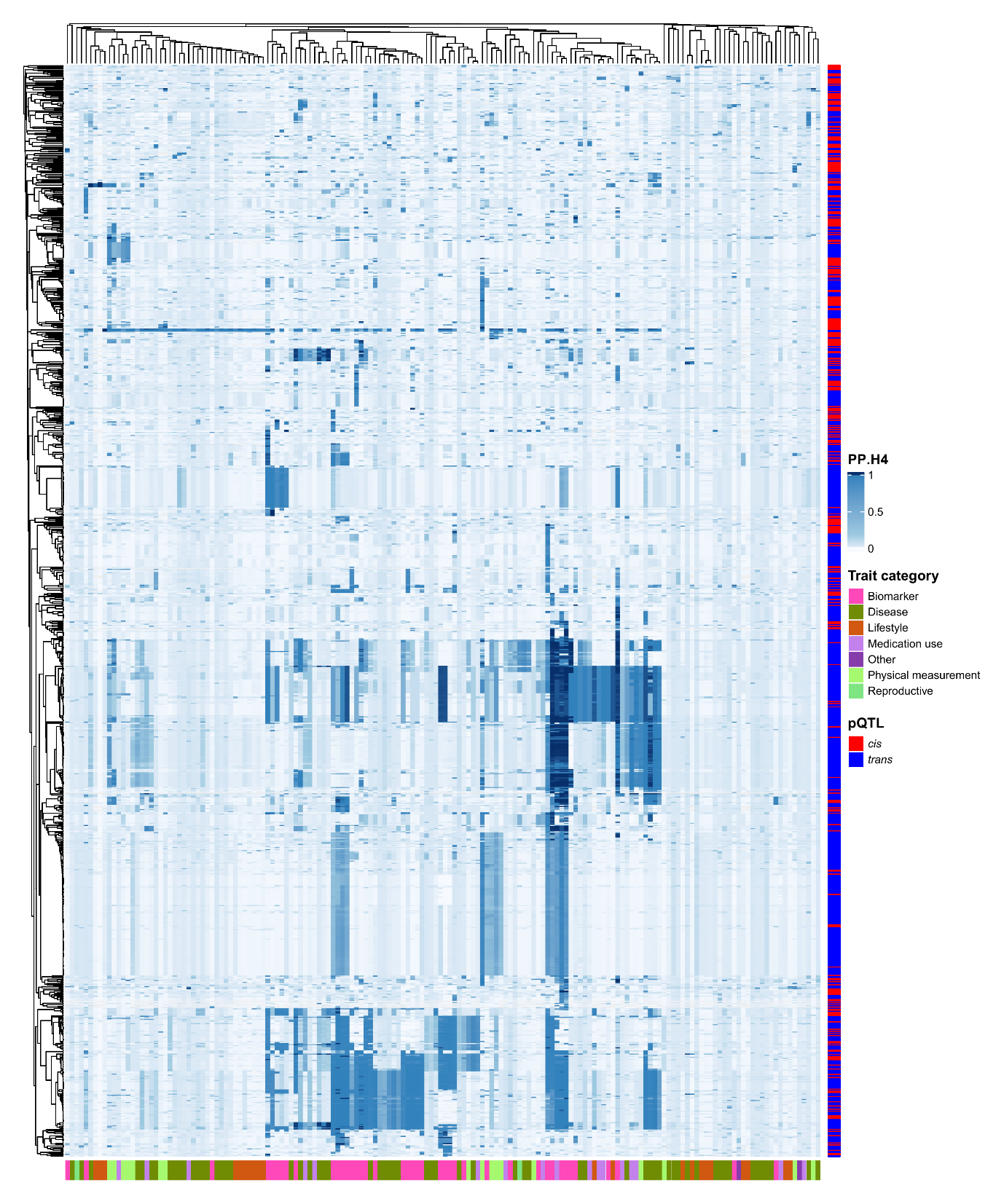

**Supplementary Figure 10. Dendrograms for trait clusters based on colocalisation posterior probabilities (PP.H4).** (A) Dendrogram for diseases and medications. (B) Dendrogram for phenotypes. (C) Combined dendrogram including diseases, medications, and phenotypes.

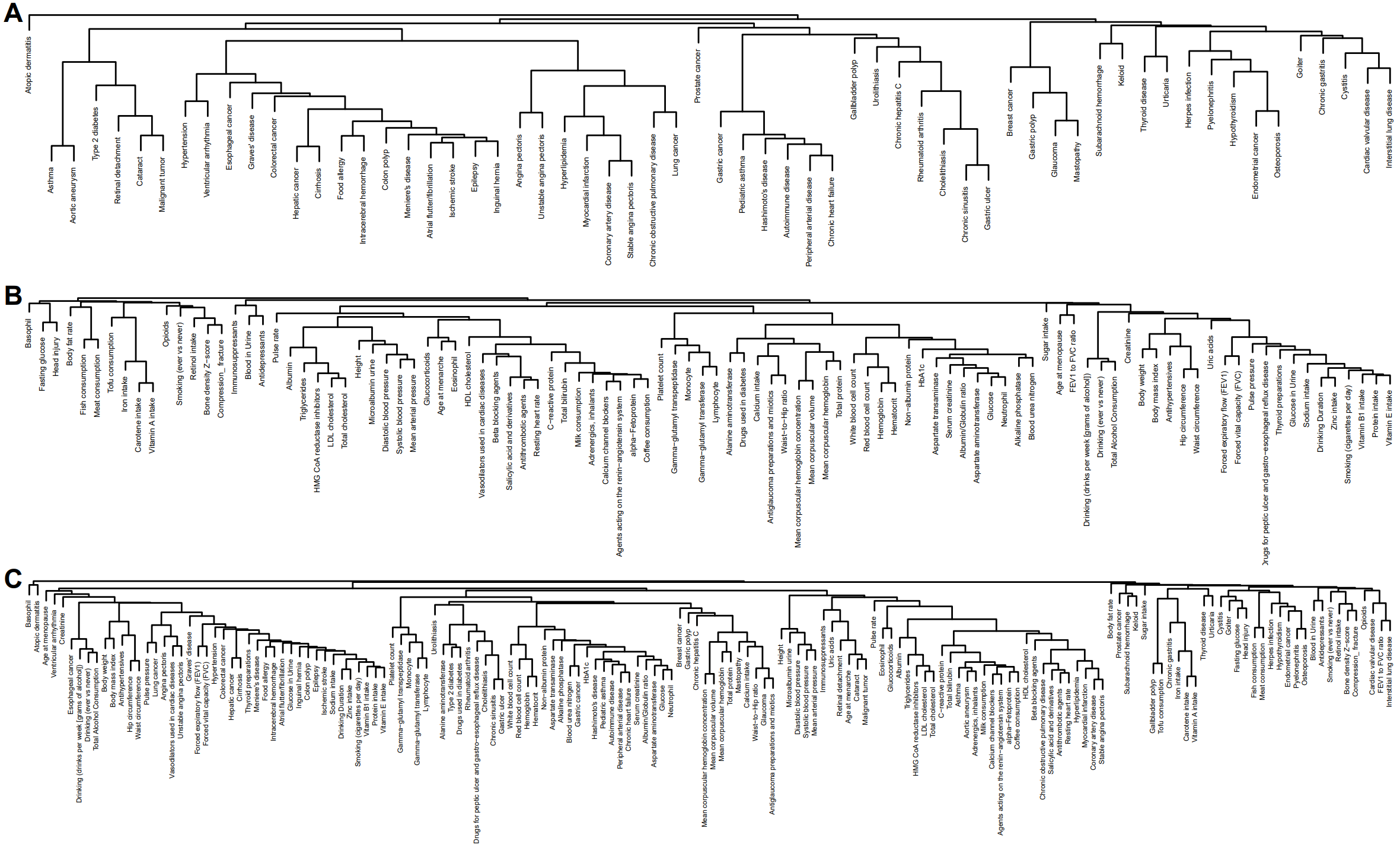

**Supplementary Figure 11. Barplots showing the top 30 traits across the three analytical approaches.** Protein counts reflect the number of proteins associated with each trait at FDR < 0.05 for PheWAS and Mendelian randomisation, and the number of colocalised proteins with PP.H4 > 0.8 for colocalisation.

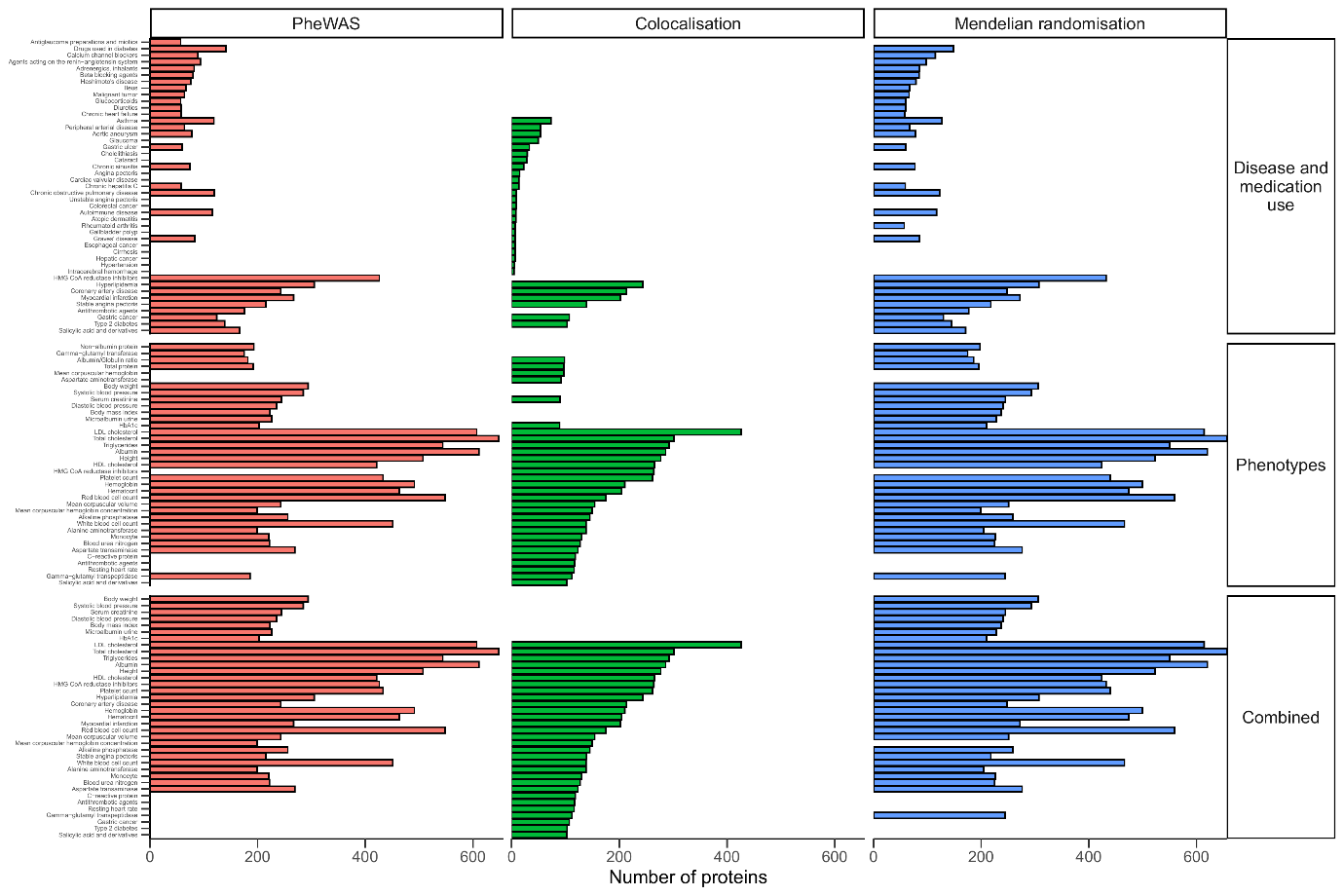

**Supplementary Figure 12. Barplots showing the top 50 proteins across the three analytical approaches.** Trait counts reflect the number of traits associated with each protein at FDR < 0.05 in PheWAS and Mendelian randomisation, and the number of traits colocalised with PP.H4 > 0.8 in colocalisation.

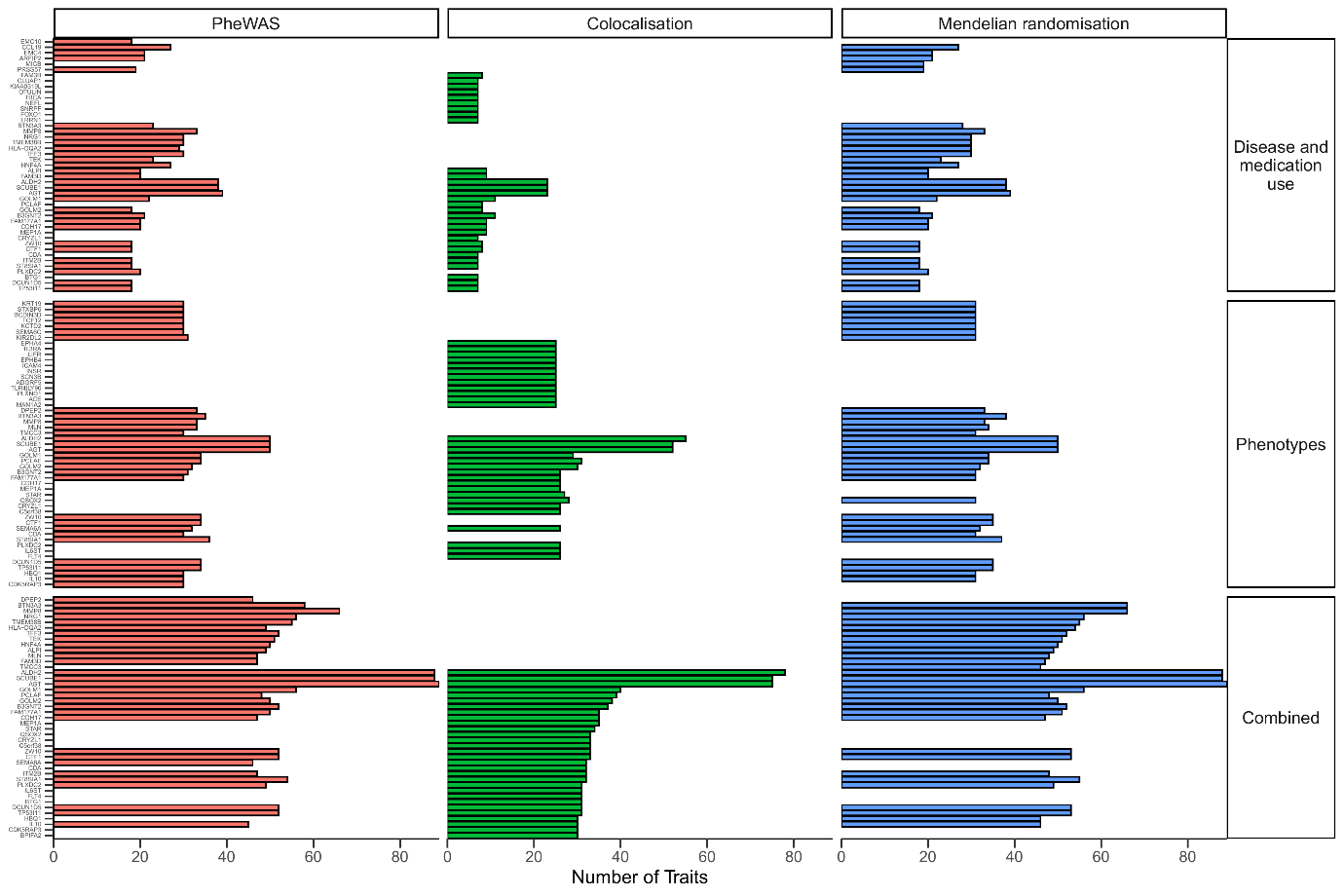
